# Evaluating the Joint Effects of Recurrent Copy Number Variants and Polygenic Scores on the Risk of Psychiatric Disorders in the iPSYCH2015 Case-Cohort Sample

**DOI:** 10.1101/2024.09.23.24314234

**Authors:** Morteza Vaez, Simone Montalbano, Ryan Waples, Morten Dybdahl Krebs, Kajsa-Lotta Georgii Hellberg, Jesper Gådin, Jonas Bybjerg-Grauholm, Preben B. Mortensen, Anders D. Børglum, Merete Nordentoft, Daniel H. Geschwind, Dorte Helenius, Thomas Werge, Andrew J. Schork, Andrés Ingason

## Abstract

The impact of rare recurrent copy number variants (rCNVs) and polygenic background attributed to common variants, on the risk of psychiatric disorders is well-established in separate studies. However, it remains unclear how polygenic background modulates the effect of rCNVs. Using the population-representative iPSYCH2015 case-cohort sample (N=96,599), we investigated the association between absolute risk of psychiatric disorders and carriage of rCNVs and polygenic scores (PGS), as well as the interaction effect between the two on disease risk. Carriers of rCNVs with higher gene constraint scores had an increased absolute risk for autism, ADHD, and schizophrenia, but not depression, whereas an increase in PGS for each respective disorder was associated with higher absolute risk across all four disorders. Similarly, elevated absolute risks were observed with the increase of both PGS and gene constraints of rCNVs except in the case of depression. In contrast to some previous case-control studies, our joint analysis of rCNV groups and PGS revealed no indication of significant interactive effect between these two factors on disease risk. Also, we found no significant interactions of PGS with any of the most common individual rCNVs, except in the case of 16p13.11 duplication, which was found to attenuate the effect of ADHD-PGS on the absolute risk of ADHD. This study advances our understanding of the interplay between rare and common important genetic risk factors for major psychiatric disorders and sheds light on the importance of population-based samples in implementing precision medicine.

## Introduction

Genetic studies have consistently shown substantial heritability for psychiatric disorders involving both common and rare variants across the human genome. ^1–3^ The ongoing advances in genome-wide association studies (GWAS) have identified many common single nucleotide polymorphisms (SNPs) associated with an increased risk of common psychiatric disorders, such as schizophrenia (SCZ), autism spectrum disorder (ASD), attention-deficit/hyperactivity disorder (ADHD), and major depressive disorder (MDD). ^4–7^ While most of the identified risk alleles for these disorders have small effect sizes (typically an associated odds ratio (OR) <1.2), the aggregated effect of these alleles (along with other common alleles with effect sizes that fall short of genome-wide significant association) can be used to derive polygenic scores (PGS) that in turn can be used to predict case status with low to moderate accuracy. ^8–10^

In addition to common SNPs, some rarer variants have also been identified that are associated with risk of psychiatric disorders. These variants include both single nucleotide variants that disrupt protein-coding sequences ^11–14^ and structural variants that either disrupt exons of single genes ^15^ (such as NRXN1) or change the dosage of entire genes through deletion or duplication of large genomic regions (such as rare recurrent copy number variants; rCNVs).^16,17^ Such rare protein-disrupting and/or gene-dosage-altering variants that have been found to associate with risk of psychiatric disorders, usually have larger effect sizes (OR>1.5) ^16^’^18^ than risk alleles of common SNPs and typically have a pleiotropic risk profile, i.e. are also associated with an increased risk of intellectual disability, epilepsy, and some somatic disorders.^18–19^

While early case-control studies indicated a very high, or even complete, penetrance of psychiatric or other neurodevelopmental disorders for some rCNVs (most prominently deletions at the velo-cardio facial syndrome (VCFS) region on 22q11)^20^, these studies often relied on case samples enriched with long-stay patients and/or patients referred to genetic analysis, and controls screened for any history of mental illness. In recent years, studies of large community- and population-based samples have challenged this perception and derived estimates indicating a more moderately increased risk associated with 22q11 deletions and other rCNVs, especially for schizophrenia.^18,21–24^ This emerging evidence of reduced penetrance of rCNVs underscores the critical role of additional etiological factors, such as polygenic background, and environmental influences, in modulating disease susceptibility.^16, 24–25^ Understanding the complex relationship between these etiological factors is important for improved diagnostic and treatment initiatives.^26^

Some earlier studies have shown the penetrance of high-risk variants to be modified by the polygenic background in both human ^27–28^ and animal disease models ^29–30^, and a few recent studies have also explored the combined effect of rCNV and PGSs on the risk of psychiatric disorders.^31–34^ For example, among 481 children affected with ADHD, Martin et al.^32^ found that carriers of any rare recurrent or non-recurrent CNV (N=60) had a significantly lower PGS for ADHD than non-carriers. Comparing the affected with 4670 control individuals, they found a corresponding significant interaction effect between carriage of CNVs and PGS with respect to risk of ADHD in a multiplicative logistic regression model.^32^ In another study, Bergen et al. evaluated the combined effects of CNVs and a PGS for schizophrenia on disease risk in a large SCZ case-control study.^33^ They observed that among affected individuals, carriers of rCNVs had significantly lower PGS for SCZ than non-carriers and that those carrying rCNVs associated with higher risk of SCZ tended to have lower PGS than those carrying rCNVs associated with less increased risk of SCZ (although, surprisingly, the same was true for controls). When fitting a model to predict case status among all rCNV carriers, they also found evidence of a significant interaction between the odds ratio for SCZ of a carried rCNV and the PGS for SCZ, in such a way that the higher the odds ratio of the carried rCNV, the less added effect of PGS was found on the prediction of case status.^33^ The results of both these studies, as well as two other studies comparing SCZ-PGS in affected carriers with unaffected controls^31^, and PGS for SCZ and intelligence quotient, and the prevalence of SCZ and intellectual disability among 22q11.2 deletion carriers^35^, are mostly consistent with a liability threshold model of disease, where PGS and CNVs contribute additively to the risk of ADHD and SCZ. In contrast to these four studies, one study found no significant difference in SCZ-PGS between affected carriers and non-carriers. ^34^

Despite these efforts, the question of how common variants modify the effects of rCNVs on the risk of psychiatric disorders remains unclear. In fact, the above-mentioned studies are conducted on case-control samples that often are biased toward cases with long-term or severe illness, and healthy controls screened for family history of mental illness. Thus, their results are not population-representative and unlikely to be generalizable to the overall population.

In this study, we benefit from the unique design of the Danish iPSYCH2015 case-cohort sample^36^ to examine the combined effect of rCNV and PGSs on the risk of psychiatric disorders in a population-representative setting. The iPSYCH2015 sample includes 140,016 individuals, consisting of a random subcohort (n=50,615) nested within a source population of all individuals who were born in Denmark between 1981-2008, and 92,531 cases encompassing all individuals from the same source population, who were diagnosed with ASD, ADHD, schizophrenia spectrum disorder (SSD), MDD or bipolar disorder (BPD) by the end of 2015. We have previously identified carriers of rCNVs at 27 genomic loci and estimated their population-based prevalence and associated risk of psychiatric disorders,^18^ and in the current study we sought to:

1. Estimate absolute risk of ASD, ADHD, SSD, and MDD associated with rCNVs and PGS until the age of 31 years, both separately and combined using survival analysis;
2. Investigate potential interactions between rCNVs and PGSs with respect to the associated risk of the four psychiatric disorders using generalized linear models (GLMs), and
3. Compare PGS of psychiatric disorders between rCNV carriers and non-carriers within cases and controls separately for the above-mentioned diagnoses.

## Results

This study analyzed data from 96,599 unrelated individuals of European ancestry (Method) within the iPSYCH2015, comprising 61,890 cases and 34,709 randomly drawn samples (i.e., sub-cohort), with an overlap of 2,292 individuals (cases that were a part of the cohort). Of the total sample, 51,507 (53%) were male and 45,092 (47%) were female. The average age at the end of follow-up was 22.01 years (mean [SD], 22.01 [7.0]), with ages ranging from 7.0 to 34.6 years. After quality control, the number of remaining verified rCNV calls totaled 2,956 at 25 rCNV loci (sTable 1). We excluded BPD from our analyses due to the small sample size.

As individual rCNVs are rare in the population (prevalence <0.5%), we needed to group rCNVs to gain sufficient power for conducting analyses of the absolute risk of ASD, ADHD, SSD, and MDD associated with rCNVs. Meanwhile, we wanted to avoid using the simple and biased approach of “any rCNV vs. none” to categorize rCNV carriers, as it has been previously demonstrated that rCNV penetrance varies across different rCNVs and outcomes.^16–18^ Hence, we considered dividing the rCNVs into various groups based on multiple approaches, including their associated odds ratios (ORs) with each diagnosis derived from iPSYCH2015^18^ and previous literature,^17,22,37–40^ their locus-wide loss-of-function observed/expected upper-bound fraction (LOEUF) score,^41^ and their pathogenicity score in ClinGen,^42^ as well as composite risk scores based on a combination of these factors (see supplementary text for details). Overall, comparisons of these grouping strategies revealed consistency in distinguishing penetrance of the different rCNV groups. Since the LOEUF score is the most objective of these measures we chose it as our primary source of grouping and divided rCNVs into three groups based on the summed LOEUF score of their contained genes (Method) as follows: rCNVs with LOEUF < 10, 10 < LOEUF < 25, and LOEUF > 25 (sFigure 1, sTable 2-6).

### Absolute Risk of ASD, ADHD, SSD, and MDD Associated with rCNV Groups in iPSYCH2015 Case-Cohort

We derived the absolute risks of ADHD, ASD, SSD, and MDD associated with each rCNV group in carriers relative to non-carriers by integrating an inverse probability weighting scheme into the survival data (Method). Overall, rCNV carriers within all three LOEUF groups had a higher absolute risk than non-carriers of being diagnosed with ASD, ADHD, and SSD, but less so for MDD (Figure 1). For ASD and ADHD, the high LOEUF group (> 25) exhibited the highest absolute risk of diagnosis, with rates of 5.5% and 6.5%, respectively, followed by the medium and low LOEUF groups, as expected from our previous study^18^. However, in the case of SSD, both the medium and high LOEUF groups displayed highly similar increases in absolute risk, at 3.8% and 4.2%, respectively. In contrast, the low LOEUF group had an increased absolute risk of SSD diagnosis by only 0.5% compared to non-carriers (Figure 1). For MDD, the observed increase in the absolute risk only amounted to 0.7-1.1% increase for any of the rCNV carrier groups compared to noncarriers (Figure 1). Having observed the rise of absolute risks with the increase in LOEUF score of the rCNV groups, we first fitted nested GLM models with and without rCNV carrier status as the independent linear covariate and then assessed the differences using likelihood ratio tests (methods). In concordance with the observations from Figure 1, the results revealed a significant increase in risk with the increase in LOEUF score levels of the rCNV groups for ASD, ADHD, and SSD (β>0.24, *P*<6.67×10^⧿10^), but not MDD (β=0.04, *P*= 0.21) (sTable 7).

**Figure 1:**
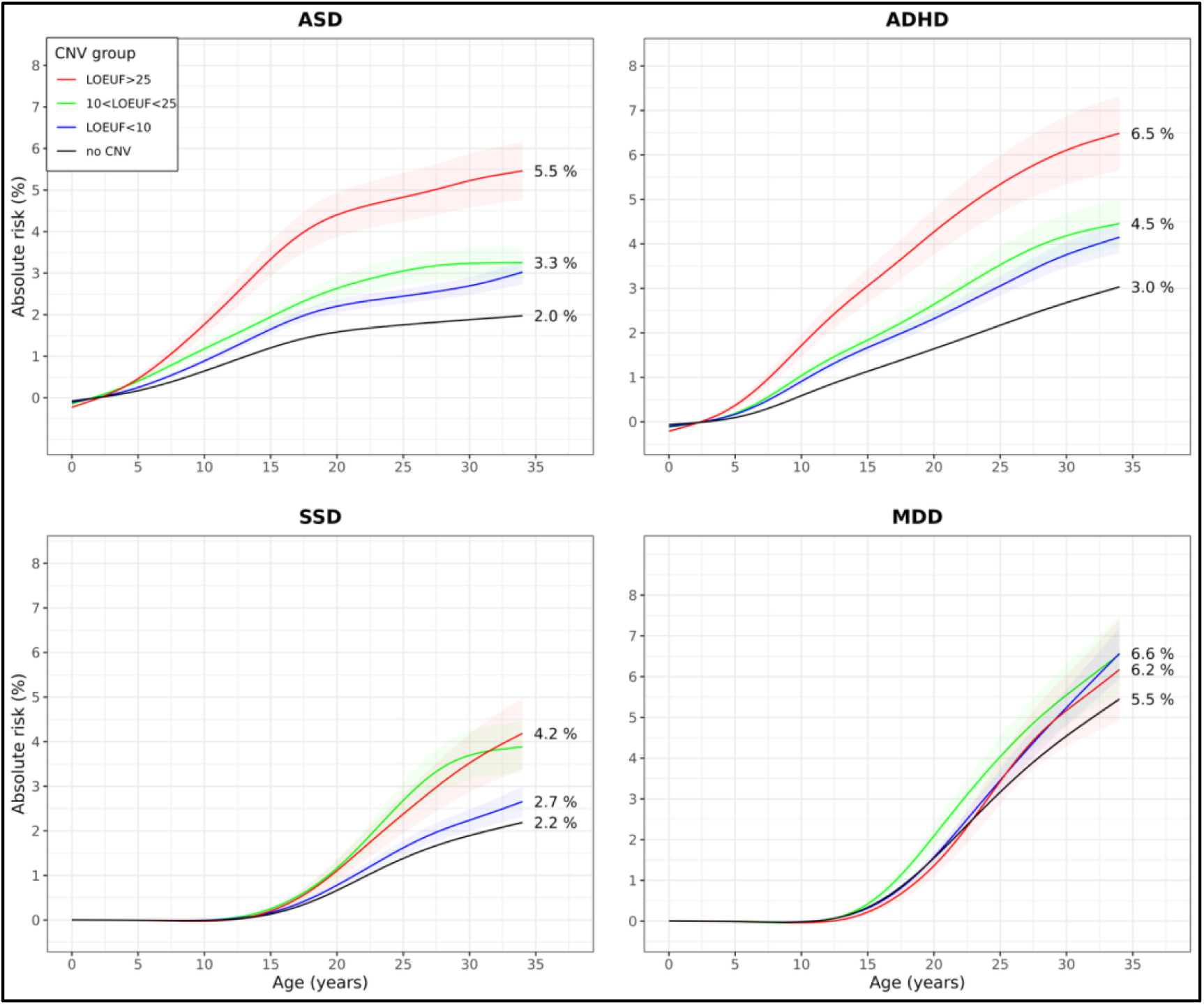
Absolute risks of ASD, ADHD, SSD, and MDD associated with rCNV groups in iPSCYH2015^36^. Absolute risks associated with rCNV groups were derived by fitting the *surv.fit* function using the survival package in R for each disorder separately. rCNVs were categorized into 3 groups based on their corresponding locus’ LOEUF score. Absolute risk curves associated with rCNVs with LOEUF below 10, LOEUF greater than 10 and lower than 25, and greater than 25, are presented by blue, green, and red colors (full), respectively. Standard errors (SE) of each rCNV group are also shown by semi-transparent colors, accordingly. All the absolute risk curves are smoothed by the *smooth.spline* function from the Spline package in R.^52^

### Absolute Risk of ASD, ADHD, SSD, and MDD associated with PGS in iPSYCH2015 Case-Cohort

We first computed the polygenic scores (PGS) of the four disorders for the entire sample using external GWASs^4^’^6^’^43^’^44^ through the SBayesR pipeline^45^. Subsequently, samples were stratified into three groups based on their PGS: low (PGS < 20th percentile), medium (20th percentile < PGS < 80th percentile), and high (PGS > 80th percentile). Absolute risk estimates for the four diagnoses were then derived in a similar way as for the rCNVs groups using the PGS group as the predictor in the survival model (methods). As expected, increased PGS was associated with an increased risk of being diagnosed with ASD, ADHD, SSD, and MDD (Figure 2). Among all groups, individuals with high PGS (> 80th percentile) exhibited the most substantial increase in absolute risk, with notable increases to 7.9% and 4.3% for MDD and ADHD, respectively (Figure 2). To formally assess the increase in the absolute risk of diagnoses associated with PGS groups, we employed GLM models similar to those in the previous step, although using PGS as the linear covariate in each model (methods). The overall results revealed that PGS was a significant positive predictor of the four diagnoses (β>0.13 and *P*>3.88×10 ^⧿45^ for each) (sTable 7).

**Figure 2:**
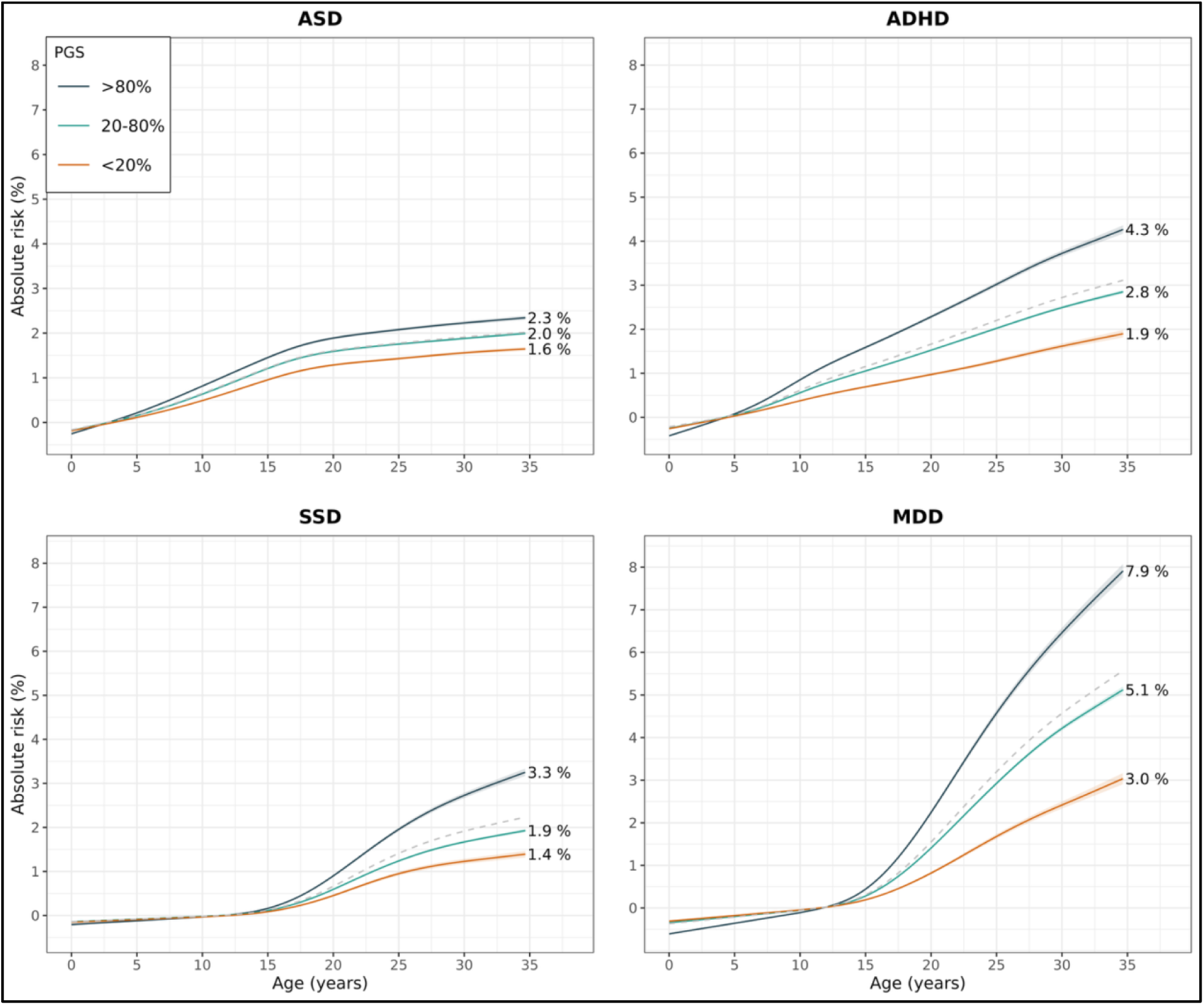
Absolute risks of ASD, ADHD, SSD, and MDD associated with polygenic score (PGS) groups in iPSCYH2015^36^. Absolute risks associated with PGS groups were derived by fitting the *surv.fit* function using the survival package^53^ in R for each disorder separately. Individuals were categorized into 3 groups based on their PGS for each corresponding disorder (i.e., PGS<20%, 20%<PGS<80%, and PGS>80%), where their associated absolute risk curves are presented by light brown, green, and dark gray colors (full), respectively. Standard errors (SE) of each PGS group are also shown by semi-transparent colors, accordingly. All the absolute risk curves are smoothed by the *smooth.spline* function from the Spline package in R.^52^

### Absolute Risk of ASD, ADHD, SSD, and MDD Associated with rCNVs and PGS Combined in iPSYCH2015 Case-Cohort

Finally, we derived the absolute risks of the four diagnoses associated jointly with both rCNVs and PGS groups using survival data, as previously described (see Methods). Overall, the absolute risk curves demonstrated that higher levels of PGS and LOEUF scores were associated with an elevated risk of diagnosis for ASD, ADHD, and SSD relative to non-carriers, with some notable exceptions within specific strata (Figure 3). In general, the absolute risk curves tended to follow the pattern observed separately for rCNV and PGS groups in additive manner. For instance, no clear distinctions in SSD diagnosis were observed between the middle and high LOEUF groups across the three PGS levels (Figure 3). Furthermore, for MDD, as previously noted, increases in PGS resulted in higher absolute risk but did not differ among carriers of the three LOEUF groups and non-carriers (Figure 3).

**Figure 3:**
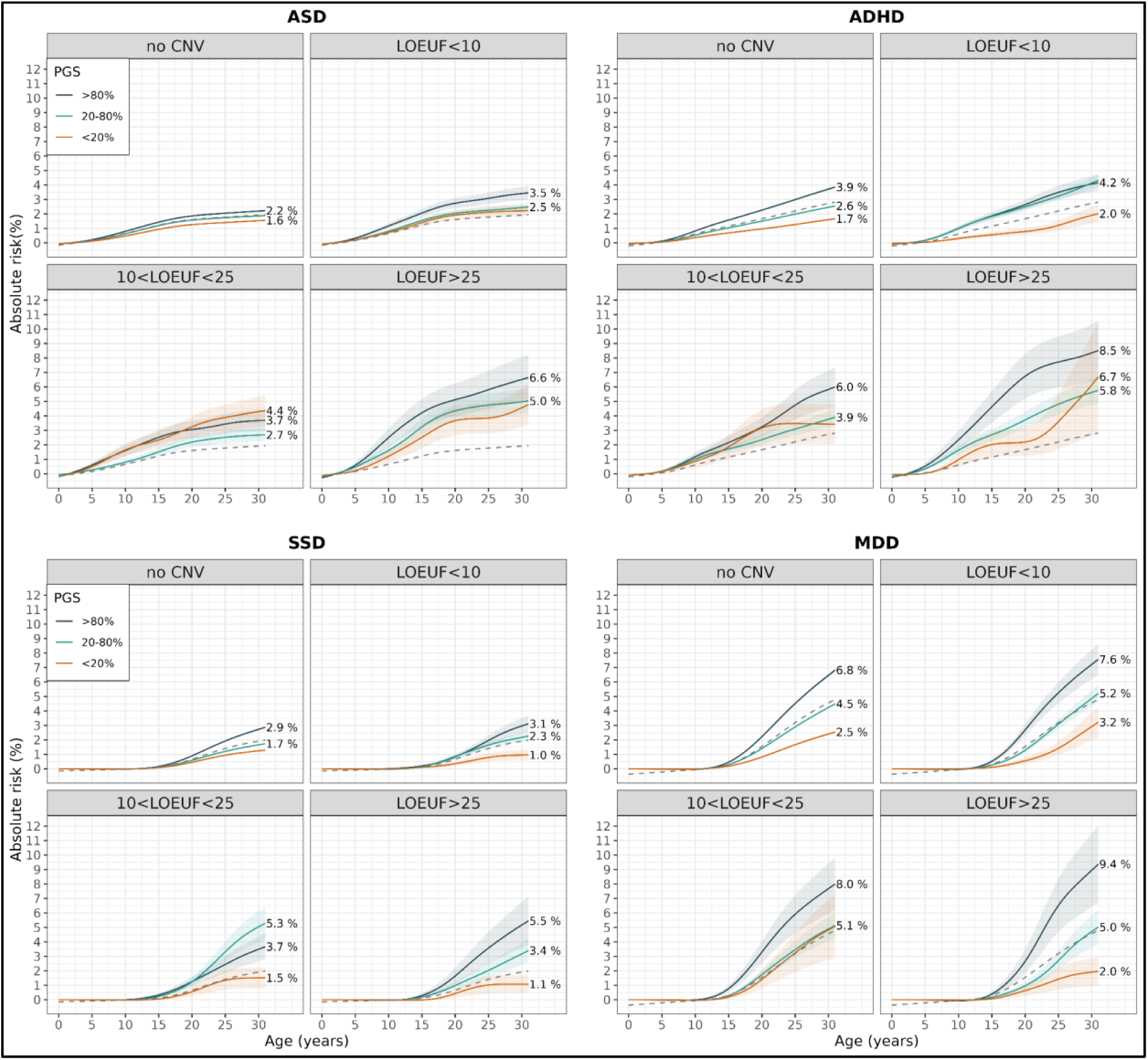
Absolute risks of ASD, ADHD, SSD, and MDD associated with rCNV and polygenic score (PGS) groups combined in ISPCYH2015^36^. Joint absolute risks associated with rCNV and PGS group were derived by fitting the *surv.fit* function using the survival package in R for each disorder separately. For enhancing the visualization, absolute risk curves associated with each disorder are shown for non-carriers, and carriers within each of rCNV groups separately in 4 different panels, stratified by the PGS groups with light brown, green, and dark gray colors. Standard errors (SE) of each PGS group are also shown by transparent colors, accordingly. Due to low carrier counts across the strata at the follow-up age above 31, absolute risk curves are shown until the follow-up age of 31. All the curves were smoothed by the *smooth.spline* function from the Spline package in R.^52^

### Model fitting results for ASD, ADHD, SSD, and MDD and investigating rCNV×PGS interactions

Following the findings from the absolute risk plots derived from the combination of rCNV and PGS groups, we investigated whether rCNVs and PGSs are associated with each diagnosis in both an additive and interactive manner using GLM models. First, we constructed the base models for predicting each diagnosis using age, sex, and origin of the sample (i.e., iPSYCH2012 and iPSYCH2015i) as the covariates and then stepwise added rCNV groups and PGS separately, and then combined (i.e., additive model) as the predictive variables. The ultimate model included the rCNV×PGS interaction as the covariate. Likelihood ratio tests were performed to evaluate the difference between each model and its corresponding nested model (Methods, sTable 8a).

The results of fitted models using only rCNVs (i.e., in the form of aggregated, rCNV groups, and rCNVs individually) were significant for ASD, ADHD, and SSD (χ^2^>38.09, *P*_LRT_>1.14×10^-5^), but not for MDD (χ^2^<23.62, *P*_LRT_>0.22). When using the PGS of each disorder to predict the corresponding disorder in the fitted models, the PGSs were positively associated with the four outcomes (χ^2^>199.75, *P*_LRT_<2.36×10^-45^). Since there were no significant associations of rCNVs with MDD, it was excluded from further additive and interactive analyses (sTable 8a).

Furthermore, the fitted additive models (i.e., rCNV + PGS) were statistically significant when including rCNVs in the aggregated manner, groups, for ASD, ADHD, and SSD (χ^2^<36.30, *P*_LRT_<6.71×10^-6^) (sTable 8a). However, the interaction analyses of rCNV×PGS (i.e., rCNVs in the form of groups and aggregated manner) across ASD, ADHD, and SSD did not show any significant results (χ^2^<3.55, *P*_LRT_>0.29) (sTable 8a & 10). As a sensitivity analysis, we repeated similar additive and interactive analyses replacing SSD with schizophrenia (SCZ) as the outcome including both rCV groups and aggregated rCNVs, in which the results consistently were significant for the additive models (χ^2^>25.41, *P*_LRT_<1.27 x 10^-5^) but not the interactive models (χ^2^<3.80, *P*_LRT_>0.14) (sTable 9).

In addition to the main analyses, we repeated all the analyses assessing the 9 most frequent rCNVs (overall carrier frequency >1∕1000) in the iPSCYH2015 dataset for ADHD, ASD, and SSD (see sTables 8b & 10-11 and method). Similarly, our findings revealed no significant rCNV×PGS interactions when assessing individual rCNVs across the three diagnoses (χ^2^<3.39, *P*_LRT_>0.06), except for the 16p13.11 duplication, which showed a significant negative interaction with PGS in predicting ADHD (χ^2^=7.66, *P*_LRT_>0.0056) (sTables 10).

We also calculated Nagelkerke’s pseudo-R² for all the fitted models to assess the explained variance. Results confirmed that the additive inclusion of rCNVs and PGS improved the explained variance for ASD, ADHD, and SSD (Figure 4, sTable 8a).

**Figure 4:**
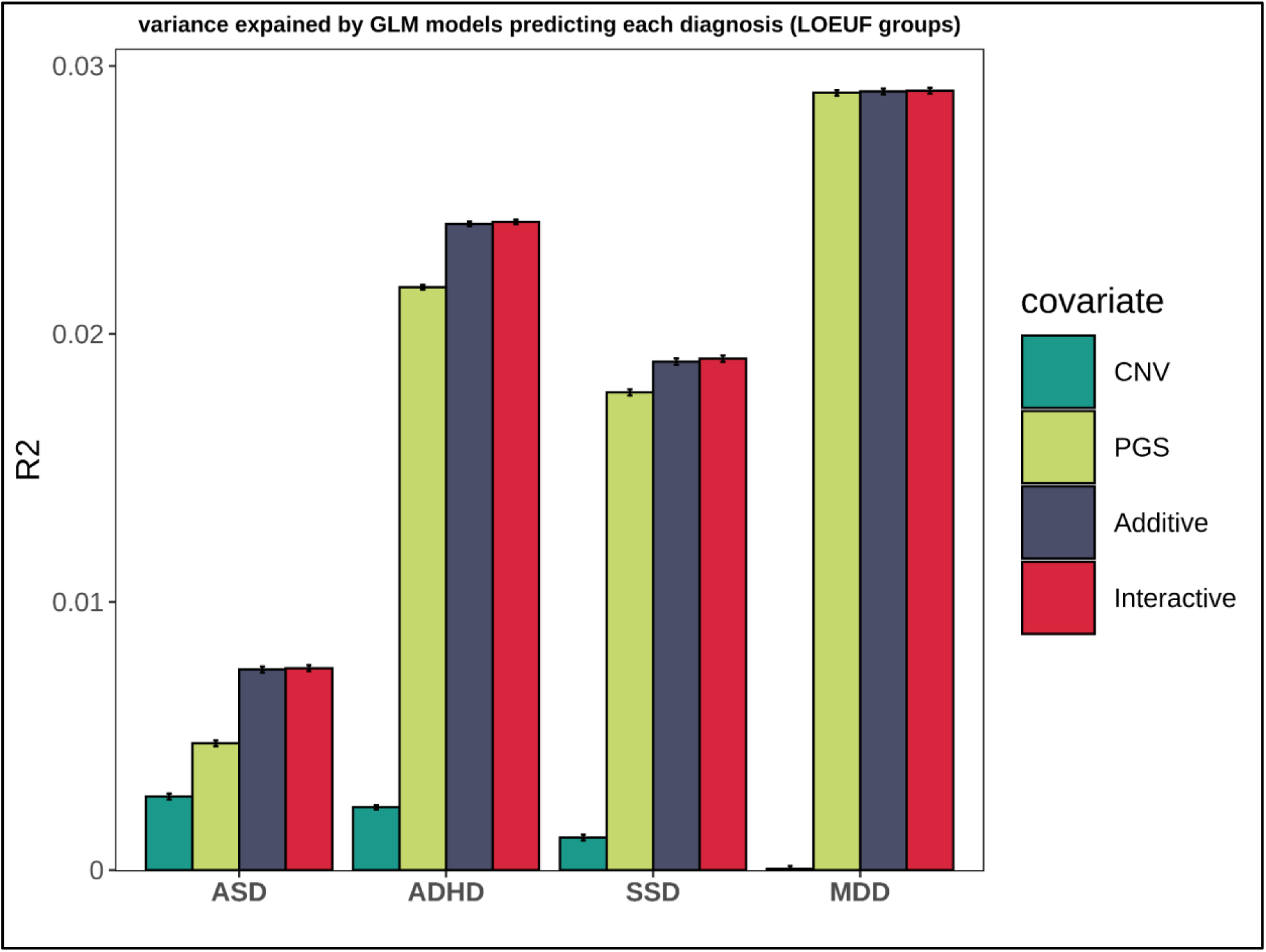
Comparison of Variance explained by fitted GLM models using rCNV groups and PGS as the explanatory genetic exposures for predicting ASD, ADHD, SSD, and MDD in iPSYCH2015. GLM models were fitted stepwise to investigate the effect of rCNV groups and PGS groups individually, additively, and interactively on each diagnosis, separately. All models included age, sex, and the genotyping array of the samples. Nagelkerke’s R-squared was computed for each model utilizing the *PseudoR2* function from the DescTools package. Each bar is colored according to the genetic covariate used in the models across the diagnoses. Dark green, light green, dark gray, and red bars indicate R2 that are derived from fitted GLMs using rCNV status, and PGS, in additive form and interactively for predicting each diagnosis, respectively. Error bars indicate standard errors (SE) of Nagelkerke’s R-squared obtained from each fitted model also were derived by bootstrapping the sample and fitting each model 1000 times (See method, sTable 5a). The results showed rCNV groups and PGS contribute significantly to the risk of ASD, ADHD, and SSD only in the additive manner and no significant interaction effects were found between rCNV groups and PGS. For MDD, there was no indication of significant rCNV effect.

However, the interactive models did not yield significant improvements in explained variance except for 16p1311 duplication in ADHD.

### Comparison of the psychiatric PGS profiles Between CNV Carriers and Non-Carriers Within Cases and Controls for ASD, ADHD, SSD, and MDD

Based on the liability threshold model, we hypothesized that carriers of rCNVs would require less additional additive risk from common variants (i.e., lower PGS) to be diagnosed with psychiatric disorders compared to non-carriers. Therefore, we compared the average PGS for each of the corresponding disorders between CNV carriers and non-carriers across rCNV groups in ADHD, ASD, SSD, and MDD cases and controls, separately (Figure 5, sTable 12) (see Methods).

**Figure 5:**
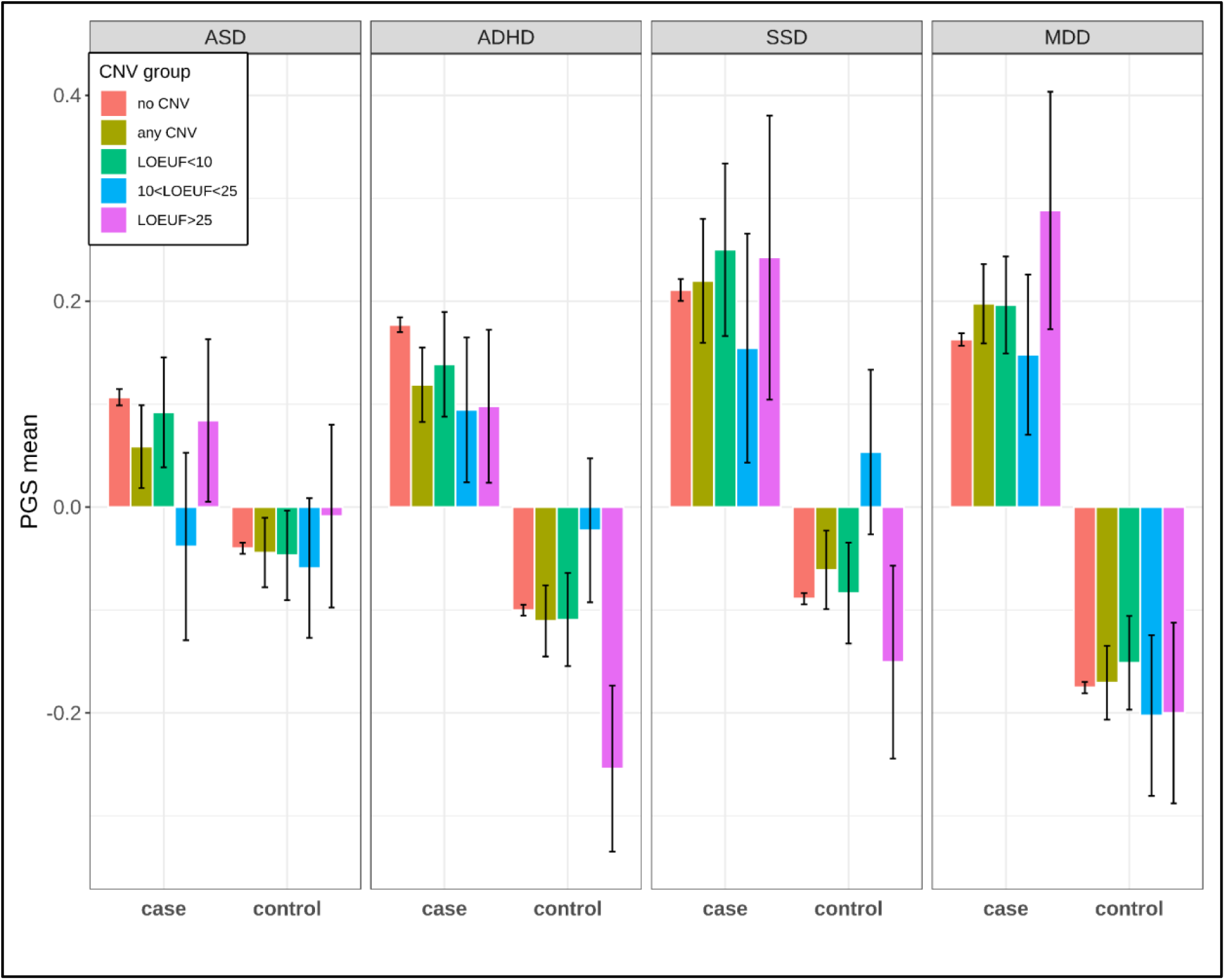
Comparison of average PGS among carriers and non-carriers across ASD, ADHD, SSD, and MDD by affection status. Mean of PGS for each diagnosis are compared among non-carriers, carriers of rCNV groups as well as carriers of any rCNV for cases and controls of ASD, ADHD, SSD, and MDD separately in different panels. Error bars indicate the SE of the PGS mean (sTable 12).

Although we observed a trend of reduced mean PGS for ASD and ADHD among cases who were rCNV carriers in the three LOEUF groups, none differed significantly from non-carriers (P_welch’s-FDR_>0.35) (Figure 5). Moreover, there was no clear trend for diminished mean PGS among SSD and MDD cases among the rCNV groups, and all tests were insignificant (P_welch’s-FDR_ >0.58). Within the controls, results were not significantly different between carriers of rCNV groups and non-carriers for each diagnosis. Similarly, the comparison of aggregated rCNV carriers with non-carriers yielded insignificant results both among cases and controls (P _welch’s FDR_>0.11). Among the three LOEUF groups, a nominally significant decrease in PGS for ADHD was observed for unaffected carriers in the highest LOEUF group compared with the middle LOEUF group, which did not remain significant after FDR correction (P_FDR_=0.35), (Figure 5, sTable 13).

## Discussion

Using 96,599 samples from the population-based iPSYCH2015 case-cohort,^36^ we investigated the absolute risk of four major psychiatric disorders—ASD, ADHD, SSD, and MDD—associated with previously identified rCNVs and common SNPs, both independently and in combination. The large sample size, comprising cases diagnosed with ASD (n=16,964), ADHD (n=20,002), SSD (n=9,749), and MDD (n=25,693), along with a random subcohort (n=37,001), provided a robust framework for evaluating the interplay between rCNVs and common SNPs concerning the risk of these psychiatric disorders. To our knowledge, this is the first study to examine the interactive effects of common and rare variants (i.e., rCNVs) on ASD, and it represents the most extensive analysis of this kind for ADHD and MDD. Our findings show that the absolute risk of ASD, ADHD, and SSD was elevated in rCNV carriers compared to non-carriers, with variations observed among different subsets of rCNVs grouped by the summed LOEUF score of the genes they affect. Similarly, an increase in PGS was associated with a higher absolute risk for all four diagnoses, and the additive effects of these two genetic predictors were confirmed for ASD, ADHD, and SSD (no rCNV-associated effect was found for the risk of MDD). Analyses of interaction between rCNVs and PGS did not reveal any significant interactive effects for ASD, ADHD and SSD, except for 16p13.11 duplication which showed a negative significant interaction with ADHD-PGS.

First, we utilized time-to-event data related to each disorder to determine the absolute risk associated with rCNVs until the age of 31 years, subdividing the rCNVs into three groups based on the summed LOEUF scores^41^ of the genes within each rCNV locus. Consistent with previous studies,^17,37,39^ we confirmed that the absolute risk for ASD, ADHD, and SSD was higher among rCNV carriers compared to non-carriers, with higher LOEUF scores of rCNVs generally indicating a greater risk than lower ones.^18^ In contrast, rCNV carriers did not exhibit an increased absolute risk of MDD, neither overall nor across groups of increasing LOEUF scores. This aligns with our previous research, which found no overall evidence for an increased rCNV-associated risk of MDD, even though this study included 9 additional rare rCNV loci.^18^ The negative MDD results differ from those of previous studies on internalizing disorders (i.e., depression and anxiety) in the UK Biobank (UKB)^22^ and two US-based healthcare community samples, ^21,46^ which reported small to moderate effects of a few rCNVs the risk of anxiety and depression. This discrepancy may be attributed to the design differences between iPSYCH2015 (its young sample and population representativeness) compared to the other samples, which has been discussed in detail elsewhere.^18^

Second, using polygenic scores as predictors in the survival analysis, we demonstrated that an increase in PGS for each diagnosis was associated with a higher absolute risk of being diagnosed with the corresponding disorder. After stratifying individuals into low-to high-risk PGS groups, we replicated the finding that PGS, indexed by the aggregated effect of SNPs, is a significant predictor for each diagnosis, particularly for ADHD, SSD, and MDD.^33,47–48,49^ However, the performance of PGS in differentiating the sample based on their PGS was suboptimal, highlighting the insufficient predictive power of current PGS and its potential utility in clinical psychiatry, which has been shown previously in other studies.^33,47–49^ Among the four assessed diagnoses, the lack of predictability power was more pronounced for PGS of ASD, with the highest PGS group exhibiting only a slight increase in the absolute risk of ASD compared to the lowest PGS group. This is in line with earlier studies that have also observed a lower predictive accuracy for ASD-PGS compared to ADHD, SSD, and MDD.^6,43,44,50^

Although the impact of both rare and common genetic variants on psychiatric disorders is well established, their combined influence, particularly in population-based samples, has not been adequately explored. Our joint analysis of rCNVs and common variants revealed that rCNVs and PGS independently influence the risk of ASD, ADHD, and SSD in an additive manner. In other words, summing the effects of both rCNVs and PGS enhances the variation explained for these disorders (but not for MDD, as discussed in the previous section). However, we found no significant interaction effects when investigating the multiplicative interaction between rCNVs and PGS to predict each diagnosis, neither when considering overall carrier status (any rCNV vs no CNV) nor when considering groups of increasing dosage effect on constrained genes. Thus, we could not confirm the hypothesis that rCNV carriers in general would require a lower PGS according to the liability threshold model to be diagnosed with psychiatric disorders than non-carriers. The comparison of the average PGS between aggregated rCNV carriers (i.e., LOEUF groups and overall carriers) and non-carriers, stratified by cases and controls for each disorder, also did not support this hypothesis, suggesting that the two risk factors combined are not sufficiently impactful to saturate a liability threshold model of risk for any of the tested disorders, at least not as estimated in this study.

For SSD, our results contrast with some previous studies.^33,35^ However, our findings from interaction analyses may not be directly comparable with the observations by studies such as Bergen et al., who reported significant interactions between odds ratios attributed to several rCNVs with SCZ-PGS among carriers of rCNVs.^33^ Also, the inconsistency between our findings and previous observations may be due to differences in study design. Clinical case-control studies (such as Bergen et al.^33^) typically include case individuals with severe and/or long-term illness, often with multiple comorbidities, and healthy controls screened for any family history of mental illness, and are therefore likely enriched with individuals representing both ends of the distribution of the genetic load of disease-relevant SNPs and rCNVs. This can result in an overestimation of the risk magnitude associated with the tested variants. For instance, our previous studies within the iPSYCH2015 case-cohort sample ^18,25,24^ have found considerably lower risk estimates for several rCNVs (most prominently the 22q11.2 deletion) for SSD than those reported in clinical case-control studies.^37,38,40^

Also for ADHD, our results contrast with those of a previous study.^32^ In this case, the failure to replicate previous observations could be attributed to differences in the study design, sample size, and/or tested rCNVs, as the previous study of Martin et al., included a much smaller case sample and only considered several large deletions.^32^ Another reason could be that the ADHD-PGS in our study is based on less well-powered summary statistics of allelic risk estimates, as ADHD cases from iPSYCH2015 constitute almost half of all cases in the most recent GWAS by the Psychiatric Genomics Consortium (PGC),^6^ and the leave-one-out summary statistics that we use are correspondingly less powered than when applying those of the full PGC-ADHD GWAS. Nonetheless, when examining the interaction of ADHD-PGS with the common individual rCNVs, we observed a significant negative interaction of the 16p13.11 duplication with PGS. This suggests an attenuation of the PGS effect on ADHD risk among carriers of this duplication, possibly due to the oversaturation of the risk among the carriers once they surpass the liability threshold. While the observed significant interaction in this rCNV may be due to distinct underlying biological processes that are currently unknown, it raises the question of why other tested rCNVs did not exhibit similar interactions. This discrepancy warrants further investigation in future studies.

For ASD, the interaction between rCNV and PGS has to our knowledge not been explored in previous studies, and we therefore have no prior studies to compare our results with. However, as with ADHD, iPSYCH2015 samples constitute a substantial part of all case samples in the largest available GWAS meta-analysis for ASD, and therefore the ASD-PGS may lack sensitivity in estimating the effect of common SNPs on ASD risk. Regarding MDD, it is unsurprising that we do not observe an interaction between the effect of common variants and rCNVS, as we do not see any effect of rCNVs on MDD risk to begin with. However, a recent study by Mollon et al.^51^ explored the impact of rCNVs and PGS of anxiety and major depressive disorder (MDD) on psychopathology scores in the UKB and discovered only a few nominally significant interactions between some individual rCNVs and PGS on the mood and anxiety scores.^51^ Consequently, the lack of strong evidence for rCNV×PGS interaction in MDD, observed in both UKB and iPSYCH, suggests that such interaction would be unlikely to be detected if investigated in other samples as well.

This study has a few limitations. First, the rarity of many rCNVs limits the power to test their interactive effects with PGS on the risk of targeted diagnoses. We tried to overcome this by grouping rCNVs by the summed constraint of the genes within each rCNV locus (summed locus LOEUF score), as we had in our previous study^18^ found evidence that increased locus LOEUF scores correlated with risk of disease, whereas the dosage change (i.e., deletion or duplication) did not. However, the LOEUF score may not optimally reflect the pathogenicity of individual rCNVs; thus, grouping rCNVs based on LOEUF score thresholds may have influenced our findings. Nonetheless, the only indication of a PGS-rCNV interaction we found, was when we looked for rCNV-specific effects among the 9 most common rCNVs, from which only duplication at 16p13.11 showed a significant interaction effect on ADHD. Second, diagnostic information was retrieved from hospital records, indicating that diagnoses made outside of the hospital setting were not accessible. Also, our use of SSD as the main measure of psychotic illness may differ from diagnostic criteria used to define schizophrenia in some previous studies; however, a sensitivity analysis of SCZ cases applying stricter criteria (ICD10:F20) found no indication of a difference from our overall results for SSD. Lastly, given the relatively young age of participants in the iPSYCH, our power to study late-onset disorders, such as bipolar disorder, was restricted. Future studies could mitigate these limitations by incorporating larger samples from broader age ranges, including diagnoses made outside of hospital settings.

## Method

### Sample and study design

iPSYCH2015^36^ is a Danish case-cohort study composed of iPSYCH2012 case-cohort and its extension; iPSYCH2015i case-cohort with 140,116 samples from the birth cohort of 1,657,449 singletons born between May 1, 1981, and December 31, 2008, who were residing in Denmark at the age of one and had a known registered mother.^36^ The iPSYCH2015 case sample comprises 92,531 psychiatric patients with inpatient or outpatient hospital diagnoses, including major depressive disorder (MDD; ICD-10: F32-F33, ICD-8: 296.09, 296.29, 298.09, 300.49, n=37,555), affective disorder (AFF; ICD-10: F30-F39, n=40,482), autism spectrum disorder (ASD; ICD-10: F84, n=24,975), bipolar disorder (BPD; ICD-10: F30-F31, n=3,819), schizophrenia spectrum disorder (SSD; ICD-10: F20-F29, n=16,008), schizophrenia (SCZ; ICD-10: F20, n=8,113), and attention-deficit/hyperactivity disorder (ADHD; ICD-10: F90, n=29,668). The cohort component includes 50,615 individuals randomly selected from the same birth cohort. All diagnostic codes and dates of diagnoses were available until December 31, 2015, which were obtained through the Psychiatric Central Research Register (PCRR)^54^.

### Genotyping, phasing, and imputation

All the individuals had their dried blood spots stored in the National Neonatal Screening Biobank (DSNB)^55^, from which the DNA samples were retrieved. Genotyping was performed on the Infinium Psych Chip v1.0 (Illumina, San Diego CA, USA) and Illumina Global Screening Array v2.0 (Illumina, San Diego CA, USA) for the individuals in iPSYCH2012 and iPSYCH2015i, respectively. Furthermore, GenomeStudio software (Illumina) was used to extract B-allele frequency and probe intensities (through log-R Ratio; LRR). Only samples with a genotyping call rate of over 95% were retained. Further details regarding the quality control and genotyping call rate are described elsewhere. ^36^

Samples genotyped on both arrays were first phased and imputed using BEAGLE5.4 separately, with HRCv1.1^56^ as the reference. Afterwards, SNPs from two separately imputed datasets were selected if passed the following criteria: 1) Genotyped at least on one of the two arrays; 2) Minimum imputation quality above 0.99 within both datasets, and 3) Maximum of 1.5% different minor allele frequency (MAF) between the two datasets. The selected SNPs from both datasets were ultimately phased and imputed again using BEAGLE5.4 aligning with HRCv1.1^56^ resulting in a total 6,518,119 SNPs with a minimum MAF>1% after performing quality control.

### Identifying genetically homogenous samples, relatedness, and PCs

We utilized a set of 20 principal components (PCs) and KING-related kinship coefficients previously obtained^57^ to identify a list of individuals of European ancestry from iPSYCH2015i, pruned for relatedness beyond the third degree. For more details, see the Supplementary text.

### CNV calling and quality control

Briefly, the selection of rCNVs in this study largely overlapped with the 54 CNVs identified in the UKB study.^19^ We excluded rCNV loci that were not listed as recurrent CNVs on ClinGen^43^ or labeled as “No Evidence” for either haploinsufficiency or triplosensitivity, resulting in 27 rCNV loci (2 of which dropped out during subsequent QC subsetting). CNV calling was performed by PennCNV^59^ and samples with an LRR-SD of 0.35 or greater, B-allele frequency drift of 0.005 or greater, and/or GC-wave factor of 0.02 or greater were removed from further analysis. PennCNV calls were validated using QCtreeCNV pipeline ^60^ providing a list of putative CNV calls, which were then visually inspected by three raters. Further details regarding the CNV calling process, quality control, and visual inspection are described thoroughly elsewhere.^18^

### CNV grouping

We evaluated whether categorizing rCNVs based on different approaches and estimating odds ratios associated with rCNV groups across the groups and approaches would result in stratification of rCNVs pathogenicity in a similar manner. The 6 grouping strategies were to group rCNVs based on; their internal odds ratios (ORs) for each disorder (i.e., rCNV-associated OR estimated previously within iPSYCH2015 data)^18^; external odds ratios (i.e., rCNV-associated odds found in the literature); simple approach of any rCNV vs no rCNV; whether rCNVs were identified to have some degrees of haploinsufficiency or triplosensitivity (HI/TS) on ClinGen^43^; LOEUF score of rCNVs; as well as composite score constructed from the three latter scores (see more details in Supplement).

### LOEUF score calculation

The LOEUF score of each rCNV locus was derived by summing the inverted LOEUF scores (1/LOEUF) of all the genes that overlapped at least 50% with the corresponding locus, obtained from the GnomADdatabase (v2.1.1)^42^ as described in our previous study.^18^ In this study, rCNV loci were categorized into three levels based on LOEUF score thresholds: <10, 10 to 25, and >25.

### Polygenic score

To compute polygenic scores, we used the latest external GWAS summary statistics of ASD^45^, ADHD^6^, SCZ^4^, and MDD^44^ obtained from the Psychiatric Genomics Consortium (PGC) after excluding iPSYCH individuals from the discovery sample). The number of SNPs used for PGS generation were 1,041,920 after intersecting SNPs in iPSYCH2015 datasets with SNPs in HapMap3 as the reference. The p-value threshold for SNPs in each GWAS was set at 0.9. SNP effects reported in each GWAS were rescaled using SBayesR^46^, based on Bayesian multiple regression with the scaling factor (γ) and the number of mixture components (π) set to the default settings (--pi 0.95,0.02,0.02,0.02,0.02,0.01; --gamma 0,0.01,0.1,1). After rescaling the SNP’s effect, PGSs were generated by applying the *--score* in *PLINKv2.00a2.3LM* to the *SBayesR’s* output files. Scores were normalized by subtracting the mean score of the control group from each individual’s score and dividing by the standard deviation of the control group’s mean score.

### Absolute Risk of ASD, ADHD, SSD, and MDD Associated with rCNV and PGS Groups in iPSYCH2015 Case-Cohort

The rCNV-associated absolute risk for each diagnosis was derived using the survfit function from the R survival package^61^, with age at first diagnosis or censoring age-depending on the event proximity-as the outcome and rCNV carrier status as the explanatory variable in each model. Censorship was implemented such that if there was a death, emigration, no diagnosis of any iPSYCH index disorder by the end of the follow-up period (December 31st, 2015), or if the participant was lost to follow-up. rCNV status was defined as a categorical variable with “no CNV” as the reference in each model. Similar models were also used when investigating absolute risk associated with PGS groups as the explanatory variable, where the lowest PGS group (i.e., PGS<20%) was the reference for this categorical variable. The definition of variables was furthermore the same when using both rCNV and PGS groups as independent variables for estimating absolute risks. All models accounted for the inverse probability of sampling (IPS) weights, following the weighting method developed by Barlow et al^62^. Due to legal restrictions in sharing individual-level data, we smoothened absolute risk curves applying *smooth.spline* function from the Spline package^53^ in R to the derived survival data in each fitted model.

To account for population stratification effects, all PGS calculations were linearly regressed against 20 principal components derived from unrelated individuals of European ancestry, and the residuals of fitted models were subsequently preserved for analysis.

### Fitting generalised linear models (GLMs)

We constructed a series of logistic regression models by GLM to investigate the effect of rCNV and PGS on the risk of each outcome, separately. All models were adjusted for age, sex, and sample origin (i.e., iPSYCH2012 or iPSYCH2015i) of subjects where the model outcome was the status for the target diagnosis coded as a binary variable. Sequentially, we introduced rCNV and PGS as predictors into each model, assessing them individually, in combination (i.e., rCNV + PGS) as well as in the interactive form of rCNV×PGS. A likelihood ratio test was employed in each step to compare the fitted model with the nested model. For each diagnosis, 54 rCNVs (i.e., corresponding to del/dup at each locus) were analyzed individually, in the form of LOEUF-groups and aggregate (i.e., any rCNV) as a categorical variable where the carrier status was labeled with regards to the reference level as having no rCNV. Furthermore, to test the effect of levels of LOEUF groups on each outcome, we fitted similar GLMs, where rCNV groups (i.e., LOEUF groups) were included as a numeric covariate after converting the LOEUF groups’ levels to 0,1,2 and 3, representing “having no rCNV”, rCNVs with “LOEUF<10”, “10<LOEUF<25” and “LOEUF>25”, respectively. When assessing the PGS effect using PGS as a linear variable as well as the categorial variable with three strata (i.e., less than 20%, between 20% and 80%, and over 80%) for all the analyses, except for the interaction analysis with the individual rCNVs. To adjust for the population stratification, all PGS scores were regressed on 20 principal components from unrelated European individuals, and the residuals were retained for further analysis. We derived the Nagelkerke’s R-squared for each model, by subtracting the R-squared of the fitted model from the R-squared of the null model utilizing the *PseudoR2* function from the DescTools package^63^. Subsequently, to compute the standard errors (SE) of Nagelkerke’s R-squared obtained from each fitted model, we first bootstrapped the sample 1000 times followed by fitting the model each time and deriving their corresponding Nagelkerke’s R-squared. We then estimated the SE of R-squared by deriving the standard deviation (SD) of the R-squareds vector corresponding to the bootstrapped models divided by the square root of 1000.

### Comparison of PGS profile between rCNV carriers and non-carriers

Welch’s t-tests were used to test the differences in the mean of PGS scores between carriers and non-carriers within cases and controls of each outcome separately.

### Ethical permissions

Research permission and access to the iPSYCH data were granted by the Danish Scientific Ethics Committee, Danish Health Authority, and the Danish Neonatal Screening Biobank Committee (https://ipsych.dk/en/data-security/health-research-and-ethicalapproval/). Following Danish legislation, constructing iPSYH data was exempted from the need for informed consent from human research participants. This study was conducted on the data stored on a secure server at the Aarhus Genome Data Center (https://genome.au.dk/).

## Data Availability

All data produced in the present study (other than sensitive person-level data, which by requirement of the data custodian and Danish legislation cannot be shared; i.e data that corresponds to fewer than 5 individuals) is available upon reasonable request to the corresponding author(s)

## Acknowledgements

This work was funded by the Tackling Multimorbidity at Scale Strategic Priorities Fund programme [grant number MR/W014416/1] delivered by the Medical Research Council and the National Institute for Health Research in partnership with the Economic and Social Research Council and in collaboration with the Engineering and Physical Sciences Research Council.

Furthermore, this study was supported by grants from the National Institutes of Health (R01-MH124789-01 and 1R01-MH124851-01), the Lundbeck Foundation (R335-2019-2318, R165-2013-15320, R155-2014-1724, R102-A9118, and R248-2017-2003), EU Horizon Europe (grant agreement 101057385 [R2D2-MH]) and the University Hospitals of Aarhus and Copenhagen. The Danish National Biobank resource was supported by the Novo Nordisk Foundation. High-performance computer capacity for handling and statistical analysis of iPSYCH data on the GenomeDK HPC facility was provided by the Center for Genomics and Personalized Medicine and the Centre for Integrative Sequencing, iSEQ, Aarhus University, Denmark

## COIs

A.D.B. has received speaker fee from Lundbeck.

