## Supplementary material for "Evaluating the Joint Effects of Recurrent Copy Number Variants and Polygenic Scores on the Risk of Psychiatric Disorders in the iPSYCH2015 Case-Cohort Sample": supplementary_material_241105.docx

### iPSYCH investigators

Anders D. Børglum^1-3^, David M. Hougaard^4^, Merete Nordentoft^5^, Ole Mors^6^, Preben B. Mortensen^2,7-9^, Thomas Werge^2,10-12^, Jakob Grove^1-3,13^, Thomas D. Als^1-3^, Alfonso Buil^10,11^, Anders Rosengren^10^, Andrés Ingason^10,11^, Andrew J. Schork^10,11^, Dorte Helenius^10^, Jesper Gådin^10^, Richard Zetterberg^10^, Vivek Appadurai^10^, Joeri Meijsen^10^, Kajsa-Lotta Georgii Hellberg^10^, Bjarni J. Vilhjálmsson^7,13^, Carsten B. Pedersen^7^, Esben Agerbo^7^, Jakob Christensen^7^, Liselotte V. Petersen^7^, Marianne Giørtz Pedersen^7^, Jonas Bybjerg-Grauholm^4^, Marie Bækvad-Hansen^4^

^1. Department of Biomedicine, Aarhus University, Aarhus, Denmark.^

^2. The Lundbeck Foundation Initiative for Integrative Psychiatric Research, iPSYCH, Denmark.^

^3. Center for Genomics and Personalized Medicine, Aarhus, Denmark.^

^4. Department for Congenital Disorders, Statens Serum Institute, Copenhagen, Denmark.^

^5. Mental Health Centre Copenhagen, Capital Region of Denmark, Copenhagen University Hospital, Copenhagen, Denmark.^

^6. Psychosis Research Unit, Aarhus University Hospital-Psychiatry, Denmark.^

^7. NCRR - National Centre for Register-Based Research, Business and Social Sciences, Aarhus University, Aarhus V, Denmark.^

^8. Centre for Integrated Register-based Research, CIRRAU, Aarhus University, Aarhus, Denmark.^

^9. Centre for Integrative Sequencing, Department of Biomedicine and iSEQ, Aarhus University, Aarhus, Denmark.^

^10. Institute of Biological Psychiatry, Mental Health Services, Copenhagen University Hospital, Copenhagen, Denmark.^

^11. Lundbeck Foundation Center for GeoGenetics, GLOBE Institute, University of Copenhagen, Copenhagen, Denmark.^

^12. Department of Clinical Medicine, University of Copenhagen, Copenhagen, Denmark.^

^13. BiRC, Bioinformatics Research Centre, Aarhus University, Aarhus, Denmark.^

### Supplementary method

**Identifying unrelated individuals of European ancestry from genotypic data:**

Ancestry information is not available for the iPSYCH individuals. Thus, to identify subjects with homogenous origin, the 1000 genomes phase 3^1^ dataset of variants was first downloaded in VCF format and multiple steps of QC were performed on the sets of variants afterward. In the QC steps, those SNPs with a minor allele frequency lower than 5%, Hardy–Weinberg p values <10^−6^, pairwise correlation (r2) >0.1 within a 1 kb region were excluded. Additionally, SNPs that did not belong to either the Infinium psych chip v1.0 or the Illumina global screening array v2.0, SNPs in regions with extended linkage disequilibrium, as well as insertion or deletions were removed from the data. Afterward, the QC-ed data was merged with both iPSYCH2012 and iPSYCH2015i datasets, and iPSYCH samples were furthermore projected into the principal components computed for 1000 genome subjects. To distinguish the samples of European and Danish ancestry, 47,586 individuals within the iPSYCH2012 sample with both Danish parents and grandparents were identified using Danish civil registers^2^, for which the first 10 principal components were then obtained. For each sample, the Mahalanobis distance was computed concerning the first 10 PCs corresponding to the previously 47,586 identified subjects. A sample was retained if the distance had a probability of more than 5.73 × 10^−7^ under a chi-square distribution. After performing genetic ancestry QC, 73,052 samples in iPSYCH2012 and 47,217 samples in iPSYCH2015i were flagged as local genetic ancestry inliers. Subsequently, kinship coefficients of the sample were estimated utilizing KING^3^, and individuals beyond the third-degree relatedness were pruned. Ultimately, 107,716 unrelated individuals in iPSYCH2015 were deemed to have European-Danish ancestry, which we used for all analyses in this study.

**CNV Grouping strategies and sensitivity analysis:**

We performed a series of sensitivity analyses to assess whether grouping individual rCNVs based on various methods into low to high-risk groups would reflect a similar pattern of distinct penetrance across the four diagnoses; namely, ASD, ADHD, SSD, and MDD. We explored and compared 5 grouping methods as follows:

1. Odds ratios in iPSYCH2015: Here, we grouped rCNVs based on their odds ratios (ORs) for each diagnosis previously estimated in iPSYCH2015^4^. For ASD, ADHD, and SSD, rCNVs were divided into three levels using the following cutoffs; OR≤1, 1<OR<2, and OR≥2, where rCNVs were labeled as “no evidence of pathogenicity”,”low risk”,” medium risk” and “high-risk”, and coded as “0”,“1”, “2” or “3”, respectively. However, for MDD, since there was no indication of significant rCNV association^4^, rCNVs were split into two groups; namely “low-risk” and high-risk” with OR of 1 set as the cut-off and coded as “1” and “2”, respectively.
2. Odds ratios in literature:^5^^-^^10^rCNVs were divided using the external ORs, wherever available. rCNVs with OR≥2 (coded as 2) for the corresponding disorder were grouped as high risk, otherwise labeled as low risk for those with OR<2 or missing risk estimate (coded as 1).
3. ClinGen^11^ haploinsufficiency/triplosensitivity index: We defined an rCNV as high risk if it was identified by the ClinGen^11^ website as an rCNV with some evidence for haploinsufficiency/triplosensitivity, else as low-risk rCNV across the 4 outcomes (i.e., low risk and high risk were labeled with score 1 and 2, respectively).
4. LOEUF score:^12^ rCNVs were split into three groups with regards to their inverse LOEUF scores(1/LOEUF) as low-LOEUF (<10, medium-LOEUF (10-25) and high-LOEUF (>25) corresponding to the arbitrary scores “1”,”2” and “3”, respectively.
5. Composite score (CS): Composite scores for each rCNV were generated by summing up their given scores within each of the three latter approaches, namely; external ORs, ClinGen, as well as lOEUF score, resulting in CS scores ranging from 3 to 7 which then were converted to 1,2,3 corresponding to low, medium and high-risk groups ultimately.

6. Any CNV vs. no CNV: Lastly, we collapsed all the rCNV carriers at 27 loci in one aggregated group.

We then calculated the risk estimates associated with the rCNV group for predicting each diagnosis using generalized linear models (GLMs) across the 6 grouping approaches. In each model, rCNV status was used as the categorical explanatory variable accounting for age, sex, and sample origin (i.e., iPSYCH2012 or iPSYCH2015i). rCNV status was defined as being rCNV carrier belonging to the CNV group, with the reference set as “non-carriers”. Only cases diagnosed with the respected outcome or controls from the random cohort entered the relevant analysis. To be consistent in grouping rCNVs across different grouping methods, We restricted the sensitivity analyses to 34 rCNVs at 18 loci that had corresponding risk estimates associated with the four diagnoses derived from iPSYCH2015.^4^

### Supplementary Figure 1

*Associated Odds ratios and confidence intervals resulted from the sensitivity analysis for grouping rCNVs across 6 different approaches.*

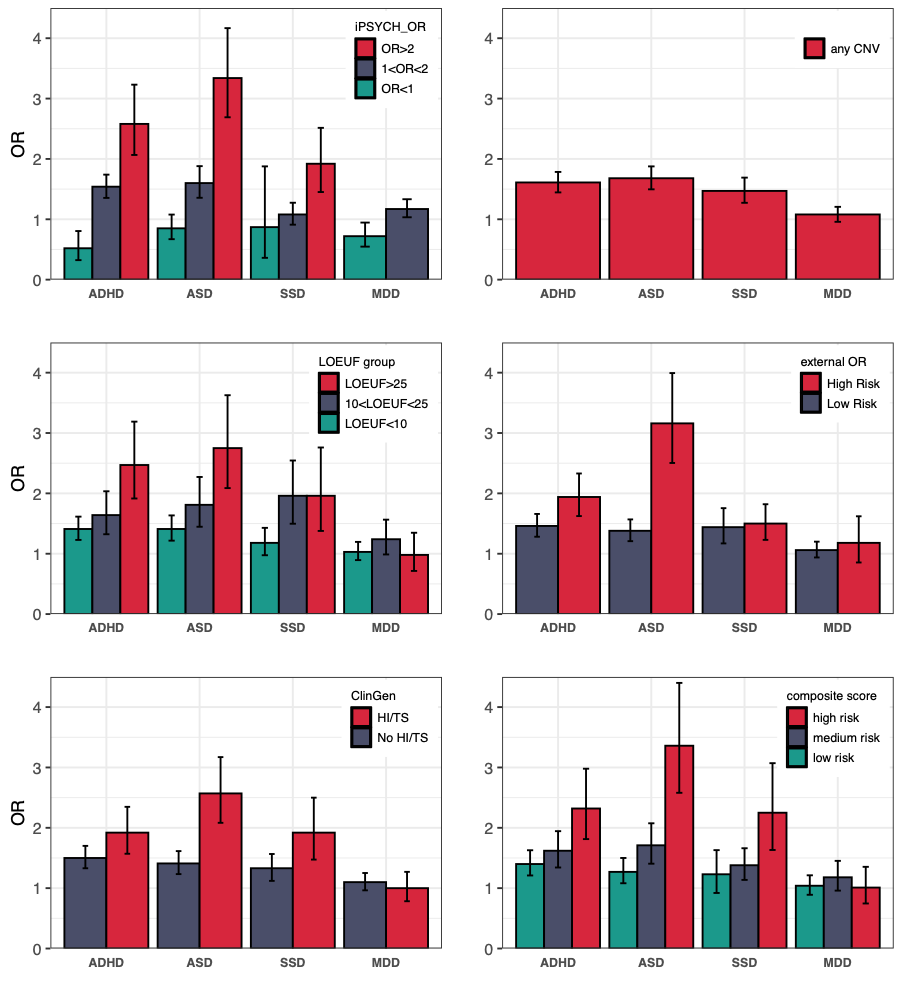

*rCNV group-associated odds ratio (ORs) and confidence intervals (CI95%) were derived from generalized linear models (GLMs) across six different grouping approaches for each diagnosis, separately. GLM results from rCNV grouping using rCNV-associated ORs in IPSYCH2015 for each disorder, any rCNV vs. no rCNV approach, rCNVs LOEUF scores, rCNV-associated external ORs for each disorder, indication of haploinsufficiency or triplosensitivity of rCNVs in ClinGen**^11^**, and generated composite scores derived from the three latter approaches (i.e., LOEUF score threshold, External ORs, and ClinGen) are shown on the upper right, upper left, middle right, middle left, lower left, and lower right, respectively (see supplementary method, sTable 10 and 11). The red, dark gray, and dark green colors of the bars represent different rCNV groups within each approach. Error bars indicate 95% CI corresponding to ORs. ASD; autism spectrum disorder, ADHD; attention-deficit hyperactivity disorder, SSD; schizophrenia spectrum disorder, MDD; major depressive disorder.*

### Supplementary Table 1

*rCNV loci used in the study and their corresponding LOUEF score group*

| **rCNV locus** | **Hg19 range (Chr:Mb)** | **LOEUF (sum^-1^)^a^** | **LOUEF group^b^** | **Filtered^c^** |
| --- | --- | --- | --- | --- |
| TAR | 1:145.39-145.81 | 22.2 | medium |  |
| 1q21.1 | 1:146.53-147.39 | 9.1 | low |  |
| 2q11.2 | 2:96.74-97.68 | 43.8 | High |  |
| 2q13 | 2:111.39-112.01 | 5.0 | low |  |
| 2q21.1 | 2:131.48-131.93 | 5.1 | low |  |
| 3q29 | 3:195.72-197.35 | 41.3 | high |  |
| WBS | 7:72.74-74.14 | 50.2 | high |  |
| 7q11.23d | 7:75.14-76.06 | 17.0 | medium | yes |
| 8p23.1 | 8:8.10-11.87 | 27.5 | high |  |
| 10q11.23 | 10:49.39-51.06 | 23.5 | medium |  |
| 10q23 | 10:82.05-88.93 | 39.9 | high |  |
| 13q12.12 | 13:23.56-24.88 | 8.0 | Low |  |
| 15q11.2 | 15:22.81-23.09 | 8.4 | low |  |
| PWAS | 15:24.82-28.39 | 19.0 | medium |  |
| 15q13.3 | 15:31.08-32.46 | 8.7 | low |  |
| 15q24 | 15:72.90-78.15 | 112.3 | high | yes |
| 16p13.11 | 16:15.51-16.29 | 19.0 | medium |  |
| 16p12.1 | 16:21.95-22.43 | 9.0 | low |  |
| 16p11.2d | 16:28.82-29.05 | 21.4 | medium |  |
| 16p11.2 | 16:29.65-30.20 | 42.9 | high |  |
| 17p12 | 17:14.14-15.43 | 6.5 | low |  |
| PLS | 17:16.81-20.21 | 76.6 | high |  |
| 17q11.2 | 17:29.12-30.27 | 26.3 | high |  |
| 17q12 | 17:34.81-36.22 | 37.1 | high |  |
| 22q11.2 | 22:18.90-20.30 | 50.4 | high |  |
| 22q11.2b | 22:20,71-21,47 | 24.2 | medium |  |
| 22q11.2d | 22:21,92-23,65 | 30.6 | high |  |

*^a^ The LOEUF score of each locus was computed by inverting the sum of the LOEUF scores of all encompassing locus genes (Since lower scores indicate higher gene constraint, scores were inverted)
^b^ We grouped rCNV loci into three groups according to their locus-LOEUF score (low; LOEUF<10, medium; 10<LOEUF<25, and high; LOEUF>25) ^c^ Loci with no carriers in the unrelated European sample were excluded.*

### Supplementary Table 2

*Grouping rCNVs into different risk groups across 6 approaches for sensitivity analysis. We categorized rCNVs using 6 different approaches to find out the most suitable strategy of grouping rCNVs in this study. Grouping approaches included using rCNVs' LOEUF scores rCNVs-associated odds ratios for each diagnosis derived from iPSYCH2015 , rCNVs-associated odds ratios for each diagnosis found in literature, ClinGen; whether rCNVs were categorized with having some level of haploinsufficiency/triplosensitivity (HI/TS) on ClinGen, anyCNV; simple approach of any rCNV vs non rCNV, and composite score; which is generated based on rCNVs' risk categories in three approaches including LOEUF score, ClinGen, and External ORs (See supplementary text)." g_LOEUF"; rCNV's LOEUF group, "iPSYCH_ASD, _ADHD,_SSD, _MDD"; rCNV's risk group based on iPSYCH's ORs, "ext_ASD,_ADHD_SSD,_MDD"; rCNV's risk group based on external ORs, ClinGen; rCNV's risk level defined by ClinGen, "cs_ASD,_SDHD,"SSD,_MDD"; rCNV's risk group based on generated composite scores. Sensitivity analysis was limited to 34 rCNVs that had corresponding associated-ORs in iPSYCH.*

*See* ***next page***

| **locus** | **locus_cnv** | **g_LOEUF** | **iPSYCH_ASD** | **iPSYCH_ADHD** | **iPSYCH_SSD** | **iPSYCH_MDD** | **ext_ASD** | **ext_ADHD** | **ext_SSD** | **ext_MDD** | **clingen** | **cs_ASD** | **cs_ADHD** | **cs_SSD** | **cs_MDD** |
| --- | --- | --- | --- | --- | --- | --- | --- | --- | --- | --- | --- | --- | --- | --- | --- |
| TAR | TAR_Deletion | 2 | Low Risk | Low Risk | Medium Risk | High Risk | Low Risk | Low Risk | Low Risk | Low Risk | HI/TS | Medium Risk | Medium Risk | Medium Risk | Medium Risk |
| TAR | TAR_Duplication | 2 | Medium Risk | Medium Risk | Medium Risk | High Risk | Low Risk | Low Risk | Low Risk | Low Risk | No HI/TS | Medium Risk | Medium Risk | Medium Risk | Medium Risk |
| 1q21.1 | 1q21.1_Deletion | 1 | High Risk | Medium Risk | Medium Risk | High Risk | Low Risk | High Risk | High Risk | Low Risk | HI/TS | Medium Risk | Medium Risk | Medium Risk | Medium Risk |
| 1q21.1 | 1q21.1_Duplication | 1 | High Risk | High Risk | High Risk | High Risk | High Risk | High Risk | High Risk | High Risk | HI/TS | Medium Risk | Medium Risk | Medium Risk | Medium Risk |
| 2q11.2 | 2q11.2_Deletion | 3 | Medium Risk | High Risk | High Risk | High Risk | Low Risk | Low Risk | High Risk | High Risk | No HI/TS | Medium Risk | Medium Risk | High Risk | High Risk |
| 2q13 | 2q13_Deletion | 1 | Low Risk | Medium Risk | Medium Risk | Low Risk | Low Risk | Low Risk | High Risk | Low Risk | No HI/TS | Low Risk | Low Risk | Medium Risk | Low Risk |
| 2q13 | 2q13_Duplication | 1 | Low Risk | Medium Risk | Medium Risk | Low Risk | Low Risk | Low Risk | Low Risk | Low Risk | No HI/TS | Low Risk | Low Risk | Low Risk | Low Risk |
| 2q21.1 | 2q21.1_Deletion | 1 | High Risk | High Risk | High Risk | Low Risk | Low Risk | Low Risk | Low Risk | Low Risk | No HI/TS | Low Risk | Low Risk | Low Risk | Low Risk |
| 2q21.1 | 2q21.1_Duplication | 1 | Low Risk | Low Risk | Low Risk | Low Risk | Low Risk | Low Risk | Low Risk | Low Risk | No HI/TS | Low Risk | Low Risk | Low Risk | Low Risk |
| 10q11.23 | 10q11.23_Deletion | 2 | Low Risk | Low Risk | Low Risk | Low Risk | Low Risk | Low Risk | Low Risk | Low Risk | No HI/TS | Medium Risk | Medium Risk | Medium Risk | Medium Risk |
| 10q11.23 | 10q11.23_Duplication | 2 | Medium Risk | Low Risk | Medium Risk | Low Risk | Low Risk | Low Risk | Low Risk | Low Risk | No HI/TS | Medium Risk | Medium Risk | Medium Risk | Medium Risk |
| 13q12.12 | 13q12.12_Deletion | 1 | Low Risk | High Risk | Low Risk | Low Risk | Low Risk | Low Risk | Low Risk | Low Risk | No HI/TS | Low Risk | Low Risk | Low Risk | Low Risk |
| 13q12.12 | 13q12.12_Duplication | 1 | High Risk | Medium Risk | High Risk | High Risk | Low Risk | Low Risk | Low Risk | Low Risk | No HI/TS | Low Risk | Low Risk | Low Risk | Low Risk |
| 15q11.2 | 15q11.2_Deletion | 1 | Medium Risk | Medium Risk | Medium Risk | High Risk | Low Risk | Low Risk | High Risk | Low Risk | No HI/TS | Low Risk | Low Risk | Medium Risk | Low Risk |
| 15q11.2 | 15q11.2_Duplication | 1 | Low Risk | Medium Risk | Medium Risk | High Risk | Low Risk | Low Risk | Low Risk | Low Risk | No HI/TS | Low Risk | Low Risk | Low Risk | Low Risk |
| PWAS | PWAS_Duplication | 2 | High Risk | High Risk | High Risk | Low Risk | High Risk | Low Risk | High Risk | High Risk | HI/TS | High Risk | Medium Risk | High Risk | High Risk |
| 15q13.3 | 15q13.3_Deletion | 1 | High Risk | High Risk | High Risk | High Risk | High Risk | High Risk | High Risk | Low Risk | No HI/TS | Medium Risk | Medium Risk | Medium Risk | Low Risk |
| 15q13.3 | 15q13.3_Duplication | 1 | Medium Risk | Medium Risk | Medium Risk | High Risk | Low Risk | Low Risk | Low Risk | Low Risk | No HI/TS | Low Risk | Low Risk | Low Risk | Low Risk |
| 16p13.11 | 16p13.11_Deletion | 2 | Medium Risk | Medium Risk | High Risk | High Risk | Low Risk | Low Risk | Low Risk | High Risk | No HI/TS | Medium Risk | Medium Risk | Medium Risk | Medium Risk |
| 16p13.11 | 16p13.11_Duplication | 2 | Medium Risk | Medium Risk | Medium Risk | High Risk | Low Risk | High Risk | High Risk | Low Risk | No HI/TS | Medium Risk | Medium Risk | Medium Risk | Medium Risk |
| 16p12.1 | 16p12.1_Deletion | 1 | Medium Risk | Medium Risk | Medium Risk | Low Risk | Low Risk | Low Risk | High Risk | Low Risk | No HI/TS | Low Risk | Low Risk | Medium Risk | Low Risk |
| 16p12.1 | 16p12.1_Duplication | 1 | Low Risk | Low Risk | Low Risk | Low Risk | Low Risk | Low Risk | Low Risk | Low Risk | No HI/TS | Low Risk | Low Risk | Low Risk | Low Risk |
| 16p11.2d | 16p11.2d_Deletion | 2 | Low Risk | Medium Risk | High Risk | High Risk | Low Risk | High Risk | High Risk | High Risk | HI/TS | Medium Risk | High Risk | High Risk | High Risk |
| 16p11.2d | 16p11.2d_Duplication | 2 | Medium Risk | Medium Risk | High Risk | High Risk | Low Risk | Low Risk | Low Risk | Low Risk | No HI/TS | Medium Risk | Medium Risk | Medium Risk | Medium Risk |
| 16p11.2 | 16p11.2_Deletion | 3 | High Risk | Low Risk | Medium Risk | Low Risk | High Risk | High Risk | Low Risk | Low Risk | HI/TS | High Risk | High Risk | High Risk | High Risk |
| 16p11.2 | 16p11.2_Duplication | 3 | High Risk | High Risk | Medium Risk | High Risk | High Risk | High Risk | High Risk | High Risk | HI/TS | High Risk | High Risk | High Risk | High Risk |
| 17p12 | 17p12_Deletion | 1 | Low Risk | Low Risk | Low Risk | Low Risk | High Risk | High Risk | High Risk | Low Risk | HI/TS | Medium Risk | Medium Risk | Medium Risk | Medium Risk |
| 17p12 | 17p12_Duplication | 1 | Low Risk | Low Risk | Medium Risk | Low Risk | Low Risk | Low Risk | Low Risk | Low Risk | HI/TS | Medium Risk | Medium Risk | Medium Risk | Medium Risk |
| 17q12 | 17q12_Deletion | 3 | High Risk | High Risk | High Risk | Low Risk | High Risk | Low Risk | High Risk | Low Risk | HI/TS | High Risk | High Risk | High Risk | High Risk |
| 17q12 | 17q12_Duplication | 3 | High Risk | High Risk | High Risk | High Risk | Low Risk | High Risk | High Risk | Low Risk | HI/TS | High Risk | High Risk | High Risk | High Risk |
| 22q11.2 | 22q11.2_Deletion | 3 | Medium Risk | Medium Risk | High Risk | Low Risk | Low Risk | High Risk | High Risk | Low Risk | HI/TS | High Risk | High Risk | High Risk | High Risk |
| 22q11.2 | 22q11.2_Duplication | 3 | High Risk | High Risk | Medium Risk | High Risk | High Risk | High Risk | Low Risk | Low Risk | HI/TS | High Risk | High Risk | High Risk | High Risk |
| 22q11.2b | 22q11.2b_Deletion | 2 | High Risk | Medium Risk | High Risk | High Risk | Low Risk | Low Risk | Low Risk | Low Risk | No HI/TS | Medium Risk | Medium Risk | Medium Risk | Medium Risk |
| 22q11.2b | 22q11.2b_Duplication | 2 | Medium Risk | High Risk | High Risk | High Risk | Low Risk | Low Risk | Low Risk | Low Risk | No HI/TS | Medium Risk | Medium Risk | Medium Risk | Medium Risk |

### Supplementary Table 3

*Association analyses of rCNV groups across different grouping approaches for ASD, ADHD, SSD, and MDD. For grouping sensitivity analysis, we computed Odds ratios (ORs) and confidence intervals asscoaited with rCNV groups across 6 different grouping approaches for each diagnosis. Grouping approaches for divinding rCNVs into different risk levels included, LOEUF; using LOEUF scores of rCNVs, iPSYCH-OR; rCNVs-asscoiatd Ors of each diagnosis in iPSYCH , anyCNV; simple appproach of having any rCNV vs no rCNV, Ext_OR; rCNVs-asscoaied ORs found in litterature for each diagnosis, ClinGen; whether rCNVs were categorized with having some level of apploinsufficiency/triplosensitivity (HI/TS) on ClinGen , and composite score; computed based on risk categroies created by LOEUF, ClinGen, and Ext_OR approaches (see Supplementary text, sFigure1,sTable10,12). "HI/TS" and No" HI/TS" are defined as whether rCNV have HI/TS or not according to ClinGen*

*Number of rCNV carriers and non carriers within cases and cotrols of each risk group are specified in the last four columns. ASD; autism spectrum disorder, ADHD; attention-deficit hyperactivity disorder, SSD; schizophrenia spectrum disorder, MDD; major depressive disorder*

| **Diagnosis** | **Risk group** | **OR** | **P** | **LCI** | **UCI** | **Grouping Strategy** | **N_carriers_case** | **N_carriers_control** | **N_noCNV_case** | **N_noCNV_ctr** |
| --- | --- | --- | --- | --- | --- | --- | --- | --- | --- | --- |
| ASD | LOEUF<10 | 1.41 | 4.89E-06 | 1.22 | 1.63 | LOEUF | 332 | 510 | 16323 | 35593 |
| ASD | 10<LOEUF<25 | 1.81 | 2.25E-07 | 1.45 | 2.27 | LOEUF | 155 | 190 | 16323 | 35593 |
| ASD | LOEUF>25 | 2.75 | 6.76E-13 | 2.09 | 3.63 | LOEUF | 126 | 107 | 16323 | 35593 |
| ADHD | LOEUF<10 | 1.41 | 8.46E-07 | 1.23 | 1.61 | LOEUF | 392 | 513 | 19271 | 35425 |
| ADHD | 10<LOEUF<25 | 1.64 | 6.39E-06 | 1.32 | 2.03 | LOEUF | 166 | 190 | 19271 | 35425 |
| ADHD | LOEUF>25 | 2.47 | 3.59E-12 | 1.92 | 3.19 | LOEUF | 147 | 111 | 19271 | 35425 |
| SSD | LOEUF<10 | 1.18 | 0.08 | 0.97 | 1.43 | LOEUF | 157 | 488 | 9436 | 33205 |
| SSD | 10<LOEUF<25 | 1.96 | 6.76E-07 | 1.50 | 2.54 | LOEUF | 95 | 177 | 9436 | 33205 |
| SSD | LOEUF>25 | 1.96 | 0.00014 | 1.38 | 2.76 | LOEUF | 56 | 100 | 9436 | 33205 |
| MDD | LOEUF<10 | 1.03 | 0.65 | 0.89 | 1.20 | LOEUF | 385 | 476 | 24997 | 32802 |
| MDD | 10<LOEUF<25 | 1.24 | 0.06 | 0.99 | 1.56 | LOEUF | 162 | 175 | 24997 | 32802 |
| MDD | LOEUF>25 | 0.98 | 0.91 | 0.71 | 1.35 | LOEUF | 80 | 98 | 24997 | 32802 |
| ASD | OR<1 | 0.85 | 0.19 | 0.67 | 1.08 | iPSYCH-OR | 108 | 261 | 16323 | 35593 |
| ASD | 1<OR<2 | 1.60 | 1.88E-08 | 1.36 | 1.88 | iPSYCH-OR | 280 | 392 | 16323 | 35593 |
| ASD | OR>2 | 3.34 | <2.2E-16 | 2.69 | 4.17 | iPSYCH-OR | 225 | 154 | 16323 | 35593 |
| ADHD | OR<1 | 0.52 | 0.0047 | 0.32 | 0.81 | iPSYCH-OR | 25 | 87 | 19271 | 35425 |
| ADHD | 1<OR<2 | 1.54 | 1.95E-11 | 1.35 | 1.74 | iPSYCH-OR | 484 | 586 | 19271 | 35425 |
| ADHD | OR>2 | 2.58 | <2.2E-16 | 2.07 | 3.23 | iPSYCH-OR | 196 | 141 | 19271 | 35425 |
| SSD | OR<1 | 0.87 | 0.74 | 0.36 | 1.88 | iPSYCH-OR | 8 | 48 | 9436 | 33205 |
| SSD | 1<OR<2 | 1.08 | 0.37 | 0.91 | 1.27 | iPSYCH-OR | 209 | 592 | 9436 | 33205 |
| SSD | OR>2 | 1.92 | 3.45E-06 | 1.45 | 2.52 | iPSYCH-OR | 91 | 125 | 9436 | 33205 |
| MDD | OR<1 | 0.72 | 0.0190 | 0.55 | 0.95 | iPSYCH-OR | 94 | 161 | 24997 | 32802 |
| MDD | 1<OR<2 | 1.17 | 0.0135 | 1.03 | 1.33 | iPSYCH-OR | 533 | 588 | 24997 | 32802 |
| ASD | anyCNV | 1.68 | <2.2E-16 | 1.50 | 1.88 | anyCNV | 613 | 807 | 16323 | 35593 |
| ADHD | anyCNV | 1.61 | <2.2E-16 | 1.45 | 1.78 | anyCNV | 705 | 814 | 19271 | 35425 |
| SSD | anyCNV | 1.47 | 1.08E-07 | 1.27 | 1.69 | anyCNV | 308 | 765 | 9436 | 33205 |
| MDD | anyCNV | 1.08 | 0.22 | 0.96 | 1.21 | anyCNV | 627 | 749 | 24997 | 32802 |
| ASD | Low Risk | 1.38 | 1.52E-06 | 1.21 | 1.57 | Ext_OR | 422 | 669 | 16323 | 35593 |
| ASD | High Risk | 3.16 | <2.2E-16 | 2.50 | 3.99 | Ext_OR | 191 | 138 | 16323 | 35593 |
| ADHD | Low Risk | 1.46 | 9.99E-09 | 1.28 | 1.66 | Ext_OR | 443 | 570 | 19271 | 35425 |
| ADHD | High Risk | 1.94 | 4.88E-13 | 1.62 | 2.33 | Ext_OR | 262 | 244 | 19271 | 35425 |
| SSD | Low Risk | 1.44 | 0.00040 | 1.17 | 1.75 | Ext_OR | 147 | 375 | 9436 | 33205 |
| SSD | High Risk | 1.50 | 5.20E-05 | 1.23 | 1.82 | Ext_OR | 161 | 390 | 9436 | 33205 |
| MDD | Low Risk | 1.06 | 0.35 | 0.94 | 1.20 | Ext_OR | 543 | 658 | 24997 | 32802 |
| MDD | High Risk | 1.18 | 0.32 | 0.85 | 1.62 | Ext_OR | 84 | 91 | 24997 | 32802 |
| ASD | No HI/TS | 1.41 | 5.25E-07 | 1.23 | 1.61 | ClinGen | 397 | 622 | 16323 | 35593 |
| ASD | HI/TS | 2.57 | <2.2E-16 | 2.08 | 3.17 | ClinGen | 216 | 185 | 16323 | 35593 |
| ADHD | No HI/TS | 1.50 | 8.05E-11 | 1.33 | 1.70 | ClinGen | 494 | 618 | 19271 | 35425 |
| ADHD | HI/TS | 1.92 | 2.01E-10 | 1.57 | 2.35 | ClinGen | 211 | 196 | 19271 | 35425 |
| SSD | No HI/TS | 1.33 | 0.0009 | 1.12 | 1.57 | ClinGen | 213 | 582 | 9436 | 33205 |
| SSD | HI/TS | 1.92 | 1.16E-06 | 1.47 | 2.50 | ClinGen | 95 | 183 | 9436 | 33205 |
| MDD | No HI/TS | 1.10 | 0.16 | 0.96 | 1.25 | ClinGen | 491 | 571 | 24997 | 32802 |
| MDD | HI/TS | 1.00 | 0.99 | 0.78 | 1.27 | ClinGen | 136 | 178 | 24997 | 32802 |
| ASD | Low Risk | 1.27 | 0.0037 | 1.08 | 1.50 | composite score | 258 | 441 | 16323 | 35593 |
| ASD | Medium Risk | 1.71 | 6.95E-08 | 1.41 | 2.07 | composite score | 205 | 261 | 16323 | 35593 |
| ASD | High Risk | 3.36 | <2.2E-16 | 2.58 | 4.40 | composite score | 150 | 105 | 16323 | 35593 |
| ADHD | Low Risk | 1.40 | 7.05E-06 | 1.21 | 1.63 | composite score | 329 | 438 | 19271 | 35425 |
| ADHD | Medium Risk | 1.62 | 3.78E-07 | 1.34 | 1.94 | composite score | 225 | 257 | 19271 | 35425 |
| ADHD | High Risk | 2.32 | 2.84E-11 | 1.81 | 2.98 | composite score | 151 | 119 | 19271 | 35425 |
| SSD | Low Risk | 1.23 | 0.15 | 0.92 | 1.63 | composite score | 70 | 211 | 9436 | 33205 |
| SSD | Medium Risk | 1.38 | 0.00093 | 1.14 | 1.66 | composite score | 167 | 440 | 9436 | 33205 |
| SSD | High Risk | 2.25 | 5.00E-07 | 1.63 | 3.07 | composite score | 71 | 114 | 9436 | 33205 |
| MDD | Low Risk | 1.04 | 0.62 | 0.89 | 1.21 | composite score | 340 | 416 | 19271 | 35425 |
| MDD | Medium Risk | 1.18 | 0.12 | 0.96 | 1.45 | composite score | 196 | 221 | 19271 | 35425 |
| MDD | High Risk | 1.01 | 0.97 | 0.75 | 1.35 | composite score | 91 | 112 | 19271 | 35425 |

### Supplementary Table 4

*Associated-ORs and 95%CI with 34 rCNVs for ASD, ADHD, SSD, and MDD in iPSYCH2015 case-cohort used for sensitivity analysis ^4.^*

| locus | CNV | OR | Pval | LCI | UCI |
| --- | --- | --- | --- | --- | --- |
| TAR | Deletion | 0.5 | 0.25 | 0.13 | 1.52 |
| TAR | Duplication | 1.61 | 0.04 | 1.02 | 2.52 |
| TAR | Deletion | 0.47 | 0.2 | 0.13 | 1.39 |
| TAR | Duplication | 1.85 | 0.0034 | 1.23 | 2.79 |
| TAR | Deletion | 1.51 | 0.5 | 0.4 | 4.76 |
| TAR | Duplication | 1.08 | 0.81 | 0.57 | 1.93 |
| TAR | Deletion | 1.07 | 0.9 | 0.35 | 3.06 |
| TAR | Duplication | 1.48 | 0.08 | 0.96 | 2.3 |
| 1q21.1 | Deletion | 2.77 | 0.0092 | 1.3 | 6.12 |
| 1q21.1 | Duplication | 4.1 | 2.80E-08 | 2.52 | 6.84 |
| 1q21.1 | Deletion | 1.99 | 0.057 | 0.99 | 4.1 |
| 1q21.1 | Duplication | 2.22 | 0.0012 | 1.37 | 3.64 |
| 1q21.1 | Deletion | 1.64 | 0.3 | 0.61 | 4.15 |
| 1q21.1 | Duplication | 2.47 | 0.0049 | 1.3 | 4.61 |
| 1q21.1 | Deletion | 1.7 | 0.19 | 0.77 | 3.87 |
| 1q21.1 | Duplication | 1.23 | 0.51 | 0.67 | 2.24 |
| 2q11.2 | Deletion | 1.38 | 0.7 | 0.25 | 6.94 |
| 2q11.2 | Deletion | 2.45 | 0.2 | 0.62 | 10.3 |
| 2q11.2 | Deletion | 8.3 | 0.021 | 1.46 | 64.14 |
| 2q11.2 | Deletion | 3.2 | 0.18 | 0.66 | 23.16 |
| 2q13 | Deletion | 0.8 | 0.59 | 0.33 | 1.76 |
| 2q13 | Duplication | 0.96 | 0.95 | 0.24 | 3.32 |
| 2q13 | Deletion | 1.24 | 0.51 | 0.65 | 2.37 |
| 2q13 | Duplication | 1.18 | 0.75 | 0.41 | 3.38 |
| 2q13 | Deletion | 1.15 | 0.77 | 0.42 | 2.85 |
| 2q13 | Duplication | 1.5 | 0.56 | 0.32 | 5.47 |
| 2q13 | Deletion | 0.93 | 0.86 | 0.42 | 1.98 |
| 2q13 | Duplication | 0.71 | 0.59 | 0.19 | 2.4 |
| 2q21.1 | Deletion | 2.16 | 0.24 | 0.6 | 8.59 |
| 2q21.1 | Duplication | 0.56 | 0.47 | 0.08 | 2.38 |
| 2q21.1 | Deletion | 2.52 | 0.15 | 0.75 | 9.75 |
| 2q21.1 | Deletion | 2.21 | 0.4 | 0.27 | 13.19 |
| 2q21.1 | Duplication | 0.51 | 0.34 | 0.11 | 1.91 |
| 10q11.23 | Deletion | 0.81 | 0.73 | 0.21 | 2.57 |
| 10q11.23 | Duplication | 1.82 | 0.45 | 0.34 | 8.74 |
| 10q11.23 | Deletion | 0.31 | 0.14 | 0.05 | 1.21 |
| 10q11.23 | Deletion | 0.62 | 0.56 | 0.09 | 2.6 |
| 10q11.23 | Duplication | 1.81 | 0.47 | 0.32 | 9.09 |
| 10q11.23 | Deletion | 0.62 | 0.41 | 0.18 | 1.87 |
| 10q11.23 | Duplication | 0.63 | 0.6 | 0.08 | 3.36 |
| 13q12.12 | Deletion | 0.86 | 0.87 | 0.11 | 4.39 |
| 13q12.12 | Duplication | 2.66 | 0.11 | 0.8 | 9.42 |
| 13q12.12 | Deletion | 3.63 | 0.025 | 1.21 | 12.12 |
| 13q12.12 | Duplication | 1.33 | 0.68 | 0.32 | 5.21 |
| 13q12.12 | Deletion | 0.48 | 0.27 | 0.12 | 1.77 |
| 13q12.12 | Duplication | 1.7 | 0.44 | 0.42 | 6.84 |
| 15q11.2 | Deletion | 1.52 | 0.00052 | 1.2 | 1.92 |
| 15q11.2 | Duplication | 0.94 | 0.62 | 0.72 | 1.21 |
| 15q11.2 | Deletion | 1.48 | 0.00053 | 1.18 | 1.84 |
| 15q11.2 | Duplication | 1.32 | 0.015 | 1.06 | 1.64 |
| 15q11.2 | Deletion | 1.03 | 0.86 | 0.74 | 1.41 |
| 15q11.2 | Duplication | 1.25 | 0.14 | 0.92 | 1.67 |
| 15q11.2 | Deletion | 1.14 | 0.29 | 0.9 | 1.44 |
| 15q11.2 | Duplication | 1.05 | 0.68 | 0.83 | 1.34 |
| PWAS | Duplication | 19.25 | 3.70E-08 | 7.52 | 65.35 |
| PWAS | Duplication | 3.06 | 0.051 | 1.03 | 10.19 |
| PWAS | Duplication | 8.03 | 0.00063 | 2.48 | 28.41 |
| PWAS | Duplication | 0.73 | 0.7 | 0.13 | 3.71 |
| 15q13.3 | Deletion | 4.64 | 0.00015 | 2.15 | 10.66 |
| 15q13.3 | Duplication | 1.51 | 0.18 | 0.81 | 2.76 |
| 15q13.3 | Deletion | 2.47 | 0.029 | 1.11 | 5.76 |
| 15q13.3 | Duplication | 1.55 | 0.11 | 0.9 | 2.65 |
| 15q13.3 | Deletion | 2.95 | 0.018 | 1.19 | 7.39 |
| 15q13.3 | Duplication | 1.52 | 0.25 | 0.72 | 3.07 |
| 15q13.3 | Deletion | 1.79 | 0.17 | 0.79 | 4.25 |
| 15q13.3 | Duplication | 1.57 | 0.11 | 0.91 | 2.74 |
| 16p13.11 | Deletion | 1.65 | 0.064 | 0.97 | 2.81 |
| 16p13.11 | Duplication | 1.76 | 0.0015 | 1.24 | 2.49 |
| 16p13.11 | Deletion | 1.99 | 0.0043 | 1.24 | 3.2 |
| 16p13.11 | Duplication | 1.98 | 2.90E-05 | 1.44 | 2.74 |
| 16p13.11 | Deletion | 2.2 | 0.011 | 1.19 | 4.03 |
| 16p13.11 | Duplication | 1.99 | 0.0011 | 1.31 | 3 |
| 16p13.11 | Deletion | 1.14 | 0.65 | 0.65 | 1.99 |
| 16p13.11 | Duplication | 1.55 | 0.016 | 1.09 | 2.22 |
| 16p12.1 | Deletion | 1.1 | 0.69 | 0.67 | 1.79 |
| 16p12.1 | Duplication | 0.94 | 0.88 | 0.41 | 2.02 |
| 16p12.1 | Deletion | 1.85 | 0.0022 | 1.25 | 2.74 |
| 16p12.1 | Duplication | 0.56 | 0.19 | 0.22 | 1.28 |
| 16p12.1 | Deletion | 1.2 | 0.55 | 0.65 | 2.12 |
| 16p12.1 | Duplication | 0.45 | 0.21 | 0.1 | 1.36 |
| 16p12.1 | Deletion | 0.77 | 0.29 | 0.48 | 1.24 |
| 16p12.1 | Duplication | 0.96 | 0.91 | 0.45 | 1.99 |
| 16p11.2d | Deletion | 0.84 | 0.73 | 0.31 | 2.11 |
| 16p11.2d | Duplication | 1.28 | 0.55 | 0.56 | 2.8 |
| 16p11.2d | Deletion | 1.03 | 0.95 | 0.45 | 2.27 |
| 16p11.2d | Duplication | 1.69 | 0.12 | 0.86 | 3.31 |
| 16p11.2d | Deletion | 2.03 | 0.12 | 0.82 | 4.89 |
| 16p11.2d | Duplication | 2.07 | 0.062 | 0.94 | 4.42 |
| 16p11.2d | Deletion | 1.01 | 0.98 | 0.42 | 2.39 |
| 16p11.2d | Duplication | 1.22 | 0.57 | 0.62 | 2.42 |
| 16p11.2 | Deletion | 3.9 | 3.20E-06 | 2.22 | 7.03 |
| 16p11.2 | Duplication | 2.52 | 5.30E-05 | 1.61 | 3.96 |
| 16p11.2 | Deletion | 0.93 | 0.84 | 0.42 | 1.92 |
| 16p11.2 | Duplication | 3.13 | 9.80E-09 | 2.13 | 4.65 |
| 16p11.2 | Deletion | 1.28 | 0.56 | 0.54 | 2.79 |
| 16p11.2 | Duplication | 1.86 | 0.024 | 1.07 | 3.16 |
| 16p11.2 | Deletion | 0.4 | 0.044 | 0.15 | 0.94 |
| 16p11.2 | Duplication | 1.16 | 0.56 | 0.71 | 1.87 |
| 17p12 | Deletion | 0.74 | 0.48 | 0.31 | 1.65 |
| 17p12 | Duplication | 0.59 | 0.45 | 0.13 | 2.12 |
| 17p12 | Deletion | 0.41 | 0.061 | 0.15 | 0.98 |
| 17p12 | Duplication | 0.71 | 0.57 | 0.19 | 2.21 |
| 17p12 | Deletion | 0.48 | 0.25 | 0.11 | 1.48 |
| 17p12 | Duplication | 1.54 | 0.48 | 0.41 | 4.85 |
| 17p12 | Deletion | 0.82 | 0.6 | 0.37 | 1.77 |
| 17p12 | Duplication | 0.68 | 0.5 | 0.22 | 2.02 |
| 17q12 | Deletion | 3.86 | 0.0056 | 1.52 | 10.6 |
| 17q12 | Duplication | 3.04 | 0.0079 | 1.35 | 7.04 |
| 17q12 | Deletion | 2.12 | 0.17 | 0.72 | 6.56 |
| 17q12 | Duplication | 3.66 | 3.00E-04 | 1.85 | 7.65 |
| 17q12 | Deletion | 3.56 | 0.083 | 0.8 | 15.12 |
| 17q12 | Duplication | 2.92 | 0.025 | 1.13 | 7.59 |
| 17q12 | Duplication | 1.78 | 0.19 | 0.75 | 4.37 |
| 22q11.2 | Deletion | 1.53 | 0.39 | 0.57 | 4.09 |
| 22q11.2 | Duplication | 2.65 | 4.30E-05 | 1.67 | 4.24 |
| 22q11.2 | Deletion | 1.48 | 0.45 | 0.53 | 4.13 |
| 22q11.2 | Duplication | 2.58 | 1.40E-05 | 1.69 | 3.99 |
| 22q11.2 | Deletion | 2.41 | 0.077 | 0.91 | 6.51 |
| 22q11.2 | Duplication | 1.46 | 0.25 | 0.75 | 2.71 |
| 22q11.2 | Deletion | 0.89 | 0.83 | 0.27 | 2.68 |
| 22q11.2 | Duplication | 1.11 | 0.7 | 0.65 | 1.89 |
| 22q11.2b | Deletion | 2.55 | 0.13 | 0.74 | 8.8 |
| 22q11.2b | Duplication | 1.12 | 0.86 | 0.29 | 3.85 |
| 22q11.2b | Deletion | 1.97 | 0.25 | 0.6 | 6.49 |
| 22q11.2b | Duplication | 2.01 | 0.19 | 0.7 | 5.92 |
| 22q11.2b | Deletion | 2.64 | 0.14 | 0.7 | 9.88 |
| 22q11.2b | Duplication | 2.75 | 0.08 | 0.85 | 8.67 |
| 22q11.2b | Deletion | 1.54 | 0.52 | 0.41 | 6.33 |
| 22q11.2b | Duplication | 1.09 | 0.88 | 0.33 | 3.44 |

### Supplementary Table 5

*Associated-ORs and 95% confidence intervals with 34 rCNVs for ASD, ADHD, SSD, and MDD reported in literature^5-10^ used for sensitivity analysis. pmid; shows the citation for the corresponding estimate.*

| Diagnosis | locus | CNV | or | LCI | UCI | pmid |
| --- | --- | --- | --- | --- | --- | --- |
| ASD | 1q21.1 | Deletion | 1.6 | 0.2 | 11.7 | 22424231 |
| ASD | 1q21.1 | Duplication | 8 | 3.5 | 18.4 | 22424231 |
| ASD | 15q11.2 | Deletion | 0.3 | 0.1 | 1.4 | 22424231 |
| ASD | PWAS | Duplication | 42.6 | 15.7 | 115.5 | 22424231 |
| ASD | 15q13.3 | Deletion | 10.8 | 3.5 | 33.1 | 22424231 |
| ASD | 16p13.11 | Duplication | 1.5 | 0.5 | 4 | 22424231 |
| ASD | 16p11.2 | Deletion | 9.5 | 5.2 | 17.4 | 22424231 |
| ASD | 16p11.2 | Duplication | 11.8 | 6.1 | 22.7 | 22424231 |
| ASD | 17p12 | Deletion | 4 | 0.9 | 17.5 | 22424231 |
| ASD | 17q12 | Deletion | 16 | 2.9 | 87.9 | 22424231 |
| ASD | 22q11.2 | Duplication | 3.3 | 1.6 | 6.6 | 22424231 |
| ADHD | 1q21.1 | Deletion | 2.68 | 0.92 | 6.44 | 31624239 |
| ADHD | 1q21.1 | Duplication | 3.44 | 1.67 | 6.52 | 31624239 |
| ADHD | 15q11.2 | Deletion | 1.65 | 1.11 | 2.37 | 31624239 |
| ADHD | 15q13.3 | Deletion | 5.97 | 2.63 | 12.6 | 31624239 |
| ADHD | 16p13.11 | Duplication | 2.12 | 1.31 | 3.27 | 31624239 |
| ADHD | 16p12.1 | Deletion | 1.52 | 0.63 | 3.16 | 31624239 |
| ADHD | 16p11.2d | Deletion | 2.19 | 0.42 | 7.29 | 31624239 |
| ADHD | 16p11.2 | Deletion | 2.16 | 0.75 | 5.11 | 31624239 |
| ADHD | 16p11.2 | Duplication | 4.34 | 2.27 | 7.81 | 31624239 |
| ADHD | 17p12 | Deletion | 2.2 | 0.67 | 5.66 | 31624239 |
| ADHD | 17q12 | Duplication | 2.2 | 0.76 | 5.24 | 31624239 |
| ADHD | 22q11.2 | Deletion | 10.73 | 4.66 | 23.15 | 31624239 |
| ADHD | 22q11.2 | Duplication | 2.24 | 1.32 | 3.63 | 31624239 |
| SSD | TAR | Deletion | 1.2 | 0.36 | 3.9 | 27602560 |
| SSD | TAR | Duplication | 1.9 | 0.93 | 3.93 | 27602560 |
| SSD | 1q21.1 | Deletion | 8.35 | 4.65 | 14.99 | 24311552 |
| SSD | 1q21.1 | Duplication | 3.45 | 1.92 | 6.2 | 24311552 |
| SSD | 2q11.2 | Deletion | 9.3 | 1.03 | 447.76 | 27602560 |
| SSD | 2q13 | Deletion | 3.6 | 0.26 | 206.03 | 27602560 |
| SSD | 2q13 | Duplication | 1.7 | 0.3 | 9.98 | 27602560 |
| SSD | 15q11.2 | Deletion | 2.15 | 1.71 | 2.68 | 24311552 |
| SSD | PWAS | Duplication | 13.2 | 3.72 | 46.77 | 24311552 |
| SSD | 15q13.3 | Deletion | 7.52 | 3.98 | 14.19 | 24311552 |
| SSD | 16p13.11 | Deletion | 1.1 | 0.43 | 2.73 | 27602560 |
| SSD | 16p13.11 | Duplication | 2.3 | 1.57 | 3.36 | 24311552 |
| SSD | 16p12.1 | Deletion | 3.3 | 1.61 | 7.05 | 27602560 |
| SSD | 16p11.2d | Deletion | 3.39 | 1.21 | 9.52 | 24311552 |
| SSD | 16p11.2d | Duplication | 1.2 | 0.44 | 3.08 | 27602560 |
| SSD | 16p11.2 | Deletion | 0.62 | 0.19 | 1.79 | 27602560 |
| SSD | 16p11.2 | Duplication | 9.4 | 4.2 | 20.9 | 27869829 |
| SSD | 17p12 | Deletion | 3.62 | 1.73 | 7.57 | 24311552 |
| SSD | 17q12 | Deletion | 6.64 | 1.78 | 24.72 | 24311552 |
| SSD | 17q12 | Duplication | 2.2 | 0.74 | 6.76 | 27602560 |
| SSD | 22q11.2 | Deletion | 67.7 | 9.4 | 492.8 | 27869829 |
| SSD | 22q11.2 | Duplication | 0.15 | 0.04 | 0.52 | 27869829 |
| MDD | TAR | Deletion | 0.95 | 0.29 | 2.29 | 30994872 |
| MDD | TAR | Duplication | 1.17 | 0.77 | 1.69 | 30994872 |
| MDD | 1q21.1 | Deletion | 1.11 | 0.46 | 2.22 | 30994872 |
| MDD | 1q21.1 | Duplication | 2.17 | 1.34 | 3.36 | 30994872 |
| MDD | 2q11.2 | Deletion | 2.34 | 0.69 | 6.02 | 30994872 |
| MDD | 2q13 | Deletion | 0.98 | 0.24 | 2.67 | 30994872 |
| MDD | 2q13 | Duplication | 1.29 | 0.49 | 2.9 | 30994872 |
| MDD | 15q11.2 | Deletion | 1.11 | 0.9 | 1.35 | 30994872 |
| MDD | PWAS | Duplication | 8.14 | 2.77 | 21.69 | 30994872 |
| MDD | 15q13.3 | Deletion | 0.77 | 0.13 | 2.52 | 30994872 |
| MDD | 16p13.11 | Deletion | 2.21 | 1.25 | 3.63 | 30994872 |
| MDD | 16p13.11 | Duplication | 0.87 | 0.63 | 1.78 | 30994872 |
| MDD | 16p12.1 | Deletion | 1.47 | 0.9 | 2.27 | 30994872 |
| MDD | 16p11.2d | Deletion | 2.23 | 0.92 | 4.63 | 30994872 |
| MDD | 16p11.2d | Duplication | 1.57 | 0.82 | 2.73 | 30994872 |
| MDD | 16p11.2 | Deletion | 1.21 | 0.54 | 2.34 | 30994872 |
| MDD | 16p11.2 | Duplication | 2.65 | 1.53 | 4.31 | 30994872 |
| MDD | 17q12 | Duplication | 1.55 | 0.69 | 3.02 | 30994872 |
| MDD | 22q11.2 | Deletion | 1.69 | 0.09 | 9.17 | 30994872 |
| MDD | 22q11.2 | Duplication | 1.72 | 1.12 | 2.53 | 30994872 |

### Supplementary Table 6

*Number of rCNV carriers within LOEUF groups and collectively across cohort and cases in iPSYCH2015 case-cohort (Unrelated European sample)*

| **LOEUF group** | **iPSYCH^a^** | **Cohort^b^** | **ASD** | **ADHD** | **SSD** | **MDD** |
| --- | --- | --- | --- | --- | --- | --- |
| Low | 1551 | 520 | 328 | 389 | 157 | 382 |
| Medium | 681 | 204 | 154 | 169 | 94 | 162 |
| High | 516 | 131 | 159 | 173 | 62 | 94 |
| Any rCNV | 2748 | 855 | 641 | 731 | 313 | 638 |

*^a^ Some individuals are diagnosed with more than one condition; therefore, the number of rCNV carriers within each group in the entire iPSYCH does not correspond to the sum of carrier counts across cases and cohort.^b^ Owing to the overlap between cases and the random cohort, some carriers within the random cohort may also belong to case samples.*

### Supplementary Table 7

*Associated effect sizes and standard errors of rCNV-LOEUF groups and PGS on ASD, ADHD, SSD, and MDD derived from fitted GLM models.*

|  | **rCNV groups^a^** | | | **PGS^b^** | | |
| --- | --- | --- | --- | --- | --- | --- |
| **Diagnosis** | **β** | **SE** | **P** | **β** | **SE** | **P** |
| ASD | 0.34 | 0.031 | <2.2 x 10^-16^ | 0.14 | 0.010 | <2.2 x 10^-16^ |
| ADHD | 0.29 | 0.030 | <2.2 x 10^-16^ | 0.28 | 0.009 | <2.2 x 10^-16^ |
| SSD | 0.25 | 0.040 | 6.67 x 10^-10^ | 0.28 | 0.012 | <2.2 x 10^-16^ |
| MDD | 0.04 | 0.034 | 0.22 | 0.35 | 0.009 | <2.2 x 10^-16^ |

*^a^ To statistically evaluate the observed increase in absolute risk of ASD, ADHD, SSD, but not MDD, associated with rCNV groups and PGS levels, we constructed different GLM models using rCNV status based on LOEUF groups as the linear explanatory variable for predicting each outcome separately (method). ^b^ Similarly, as we observed that an increase in PGS levels was associated with an elevated absolute risk of ADHD, ASD, SSD, and MDD, we built GLM models using PGS as the linear independent variable to predict each outcome separately. β, SE, and P correspond to the beta, standard error, and P value of the explanatory variable, respectively, derived from the summary of fitted GLM model.*

### Supplementary Table 8a

*Model fitting results for ASD, ADHD, SSD, and MDD as a function of PGS and rCNV status (as LOEUF groups and aggregated)*

| **ASD** | | | | | | **ADHD** | | | | |
| --- | --- | --- | --- | --- | --- | --- | --- | --- | --- | --- |
| **Model** | **Genetic exposure ^a^** | **df** | **Chisq ^b^** | **P ^c^** | **ΔR^2 d^** | **Model** | **df** | **Chisq ^b^** | **P ^c^** | **ΔR^2 d^** |
| 0 | None | 3 |  |  |  | 0 | 3 |  |  |  |
| 1 | CNV_groups | 4 | 116.09 | <2.2 x 10^-16^ | 0.003 | 1 | 4 | 101.4 | <2.2 x 10^-16^ | 0.002 |
| 2 | anyCNV | 4 | 88.58 | <2.2 x 10^-16^ | 0.002 | 2 | 4 | 83.14 | <2.2 x 10^-16^ | 0.002 |
| 3 | PGS | 4 | 199.99 | <2.2 x 10^-16^ | 0.005 | 3 | 4 | 945.19 | <2.2 x 10^-16^ | 0.022 |
| 4 | PGS + CNV_groups | 5 | 116.76 | <2.2 x 10^-16^ | 0.007 | 4 | 5 | 103.78 | <2.2 x 10^-16^ | 0.024 |
| 5 | PGS + anyCNV | 5 | 200.85 | <2.2 x 10^-16^ | 0.007 | 5 | 5 | 946.24 | <2.2 x 10^-16^ | 0.024 |
| 6 | PGS×CNV_groups | 8 | 2.22 | 0.52 | 0.007 | 6 | 8 | 3.3 | 0.34 | 0.024 |
| 7 | PGS×anyCNV | 6 | 0.5 | 0.48 | 0.006 | 7 | 6 | 1.12 | 0.29 | 0.024 |

| **SSD** | | | | | | **MDD ^e^** | | | | |
| --- | --- | --- | --- | --- | --- | --- | --- | --- | --- | --- |
| **Model** | **Genetic exposure ^a^** | **df** | **Chisq ^b^** | **P ^c^** | **ΔR^2 d^** | **Model** | **df** | **Chisq ^a^** | **P ^c^** | **ΔR^2 d^** |
| 0 | None | 3 |  |  |  | 0 | 3 |  |  |  |
| 1 | CNV_groups | 4 | 38.09 | 2.70 x 10^-8^ | 0.0012 | 1 | 4 | 2.76 | 0.43 | 5 x 10^-5^ |
| 2 | anyCNV | 4 | 27.40 | 1.65 x 10^-7^ | 0.0009 | 2 | 4 | 1.48 | 0.22 | 3 x 10^-5^ |
| 3 | PGS | 4 | 560.36 | <2.2 x 10^-16^ | 0.0178 | 3 | 4 | 1484.8 | <2.2 x 10^-16^ | 0.0290 |
| 4 | PGS + CNV_groups | 5 | 36.30 | 6.47 x 10^-8^ | 0.0190 | 4 |  |  |  |  |
| 5 | PGS + anyCNV | 5 | 558.40 | <2.2 x 10^-16^ | 0.0186 | 5 |  |  |  |  |
| 6 | PGS×CNV_groups | 8 | 3.55 | 0.31 | 0.0191 | 6 |  |  |  |  |
| 7 | PGS×anyCNV | 6 | 0.90 | 0.34 | 0.0186 | 7 |  |  |  |  |

*We fitted different generalized linear models (GLMs) to assess the combined and interactive effects of rCNV and PGS on ASD, ADHD, SSD, and MDD using rCNVs divided into three LOEUF groups and rCNV in an aggregated form (i.e., any rCNV vs. no rCNV) (method). Likelihood ratio tests were used to compare each model with its nested model. The null model included sex, age at the end of follow-up, and the genotyping array as the independent variables. ^a^ rCNV and PGS were added stepwise to the null model. ^b^ Chisq and ^c^ P are derived from the likelihood ratio tests. ^d^ Niekerk’s R2 was calculated for each model and ΔR^2^ was derived by subtracting R2 of each full model from R2 of the nested model. ^e^ Since no significant effect of rCNV was found on the risk of MDD, we did not report the results of the additive and interactive effect of rCNV with PGS in this case.*

### Supplementary Table 8b

*Model fitting results for ASD, ADHD, SSD, and MDD as a function of*

*PGS and rCNV status in carriers of common rCNVs in iPSYCH2015. In addition to the main*

*analyses we assessed the joint effect of rCNVs and PGS on ASD, ADHD, SSD, and MDD using*

*most common rCNVs (i.e., 1/1000 carriers in iPSYCH2015) indivdiually by adding each*

*variables step-wise by Generalised linear models (GLMs). Each model then was tested against*

*its nested model using Likelihood ratio test. rCNV and PGS collumns reffer to the rCNV and PGS that was used in each model. P and chisq values are derived from the LRT. All models were adjusted for age, sex and genotyping array. Additive and interactive models; correpsond to the models inclduing "CNV+PGS" and "CNVxPSG" as explnatpry variables, respectively. Since there was no indiction of signifgicant effect of rCNVs on MDD in "cnv_only" based models, we did not report the LRT results derived from additive or interactive models in MDD. ADHD; attention-deficit hyperactivity disorder, ASD; autism spectrum disorder, MDD; major depressive disorder, SSD; schizophrenia spectrum disorder.*

| **rCNV** | **pheno** | **pgs** | **p** | **chisq** | **df** | **model** |
| --- | --- | --- | --- | --- | --- | --- |
| TAR_dup | ASD | pgs_asd | 0.18 | 1.77 | 4 | cnv_only |
| TAR_dup | ASD | pgs_asd | <2.2E-16 | 199.99 | 4 | pgs_only |
| TAR_dup | ASD | pgs_asd | 0.16 | 2 | 5 | additive |
| TAR_dup | ASD | pgs_asd | 0.60 | 0.27 | 6 | interactive |
| TAR_dup | ADHD | pgs_adhd | 0.0094 | 6.74 | 4 | cnv_only |
| TAR_dup | ADHD | pgs_adhd | <2.2E-16 | 945.19 | 4 | pgs_only |
| TAR_dup | ADHD | pgs_adhd | 0.01 | 6.58 | 5 | additive |
| TAR_dup | ADHD | pgs_adhd | 0.91 | 0.01 | 6 | interactive |
| TAR_dup | SSD | pgs_scz | 0.91 | 0.01 | 4 | cnv_only |
| TAR_dup | SSD | pgs_scz | <2.2E-16 | 560.36 | 4 | pgs_only |
| TAR_dup | SSD | pgs_scz | 0.87 | 0.03 | 5 | additive |
| TAR_dup | SSD | pgs_scz | 0.24 | 1.35 | 6 | interactive |
| TAR_dup | MDD | pgs_mdd | 0.24 | 1.4 | 4 | cnv_only |
| TAR_dup | MDD | pgs_mdd | <2.2E-16 | 1484.8 | 4 | pgs_only |
| 1q21.1_dup | ASD | pgs_asd | 3.15E-07 | 26.16 | 4 | cnv_only |
| 1q21.1_dup | ASD | pgs_asd | <2.2E-16 | 199.99 | 4 | pgs_only |
| 1q21.1_dup | ASD | pgs_asd | 3.43E-07 | 25.99 | 5 | additive |
| 1q21.1_dup | ASD | pgs_asd | 0.86 | 0.03 | 6 | interactive |
| 1q21.1_dup | ADHD | pgs_adhd | 0.0056 | 7.67 | 4 | cnv_only |
| 1q21.1_dup | ADHD | pgs_adhd | <2.2E-16 | 945.19 | 4 | pgs_only |
| 1q21.1_dup | ADHD | pgs_adhd | 0.0074 | 7.17 | 5 | additive |
| 1q21.1_dup | ADHD | pgs_adhd | 0.32 | 0.96 | 6 | interactive |
| 1q21.1_dup | SSD | pgs_scz | 0.039 | 4.28 | 4 | cnv_only |
| 1q21.1_dup | SSD | pgs_scz | <2.2E-16 | 560.36 | 4 | pgs_only |
| 1q21.1_dup | SSD | pgs_scz | 0.049 | 3.88 | 5 | additive |
| 1q21.1_dup | SSD | pgs_scz | 0.96 | 0 | 6 | interactive |
| 1q21.1_dup | MDD | pgs_mdd | 0.86 | 0.03 | 4 | cnv_only |
| 1q21.1_dup | MDD | pgs_mdd | <2.2E-16 | 1484.8 | 4 | pgs_only |
| 15q11.2_del | ASD | pgs_asd | 0.00058 | 11.82 | 4 | cnv_only |
| 15q11.2_del | ASD | pgs_asd | <2.2E-16 | 199.99 | 4 | pgs_only |
| 15q11.2_del | ASD | pgs_asd | 0.00048 | 12.18 | 5 | additive |
| 15q11.2_del | ASD | pgs_asd | 0.95 | 0 | 6 | interactive |
| 15q11.2_del | ADHD | pgs_adhd | 0.0025 | 9.15 | 4 | cnv_only |
| 15q11.2_del | ADHD | pgs_adhd | <2.2E-16 | 945.19 | 4 | pgs_only |
| 15q11.2_del | ADHD | pgs_adhd | 0.0039 | 8.31 | 5 | additive |
| 15q11.2_del | ADHD | pgs_adhd | 0.46 | 0.54 | 6 | interactive |
| 15q11.2_del | SSD | pgs_scz | 0.46 | 0.55 | 4 | cnv_only |
| 15q11.2_del | SSD | pgs_scz | <2.2E-16 | 560.36 | 4 | pgs_only |
| 15q11.2_del | SSD | pgs_scz | 0.39 | 0.74 | 5 | additive |
| 15q11.2_del | SSD | pgs_scz | 0.71 | 0.14 | 6 | interactive |
| 15q11.2_del | MDD | pgs_mdd | 0.32 | 0.97 | 4 | cnv_only |
| 15q11.2_del | MDD | pgs_mdd | <2.2E-16 | 1484.8 | 4 | pgs_only |
| 15q11.2_dup | ASD | pgs_asd | 0.82 | 0.05 | 4 | cnv_only |
| 15q11.2_dup | ASD | pgs_asd | <2.2E-16 | 199.99 | 4 | pgs_only |
| 15q11.2_dup | ASD | pgs_asd | 0.83 | 0.05 | 5 | additive |
| 15q11.2_dup | ASD | pgs_asd | 0.26 | 1.24 | 6 | interactive |
| 15q11.2_dup | ADHD | pgs_adhd | 0.027 | 4.86 | 4 | cnv_only |
| 15q11.2_dup | ADHD | pgs_adhd | <2.2E-16 | 945.19 | 4 | pgs_only |
| 15q11.2_dup | ADHD | pgs_adhd | 0.023 | 5.2 | 5 | additive |
| 15q11.2_dup | ADHD | pgs_adhd | 0.59 | 0.3 | 6 | interactive |
| 15q11.2_dup | SSD | pgs_scz | 0.28 | 1.18 | 4 | cnv_only |
| 15q11.2_dup | SSD | pgs_scz | <2.2E-16 | 560.36 | 4 | pgs_only |
| 15q11.2_dup | SSD | pgs_scz | 0.44 | 0.6 | 5 | additive |
| 15q11.2_dup | SSD | pgs_scz | 0.84 | 0.04 | 6 | interactive |
| 15q11.2_dup | MDD | pgs_mdd | 0.68 | 0.17 | 4 | cnv_only |
| 15q11.2_dup | MDD | pgs_mdd | <2.2E-16 | 1484.8 | 4 | pgs_only |
| 16p13.11_del | ASD | pgs_asd | 0.021 | 5.33 | 4 | cnv_only |
| 16p13.11_del | ASD | pgs_asd | <2.2E-16 | 199.99 | 4 | pgs_only |
| 16p13.11_del | ASD | pgs_asd | 0.023 | 5.17 | 5 | additive |
| 16p13.11_del | ASD | pgs_asd | 0.73 | 0.11 | 6 | interactive |
| 16p13.11_del | ADHD | pgs_adhd | 0.002 | 9.46 | 4 | cnv_only |
| 16p13.11_del | ADHD | pgs_adhd | <2.2E-16 | 945.19 | 4 | pgs_only |
| 16p13.11_del | ADHD | pgs_adhd | 0.0019 | 9.7 | 5 | additive |
| 16p13.11_del | ADHD | pgs_adhd | 0.41 | 0.68 | 6 | interactive |
| 16p13.11_del | SSD | pgs_scz | 0.0016 | 9.98 | 4 | cnv_only |
| 16p13.11_del | SSD | pgs_scz | <2.2E-16 | 560.36 | 4 | pgs_only |
| 16p13.11_del | SSD | pgs_scz | 0.0011 | 10.58 | 5 | additive |
| 16p13.11_del | SSD | pgs_scz | 0.15 | 2.04 | 6 | interactive |
| 16p13.11_del | MDD | pgs_mdd | 0.87 | 0.03 | 4 | cnv_only |
| 16p13.11_del | MDD | pgs_mdd | <2.2E-16 | 1484.8 | 4 | pgs_only |
| 16p13.11_dup | ASD | pgs_asd | 0.0034 | 8.6 | 4 | cnv_only |
| 16p13.11_dup | ASD | pgs_asd | <2.2E-16 | 199.99 | 4 | pgs_only |
| 16p13.11_dup | ASD | pgs_asd | 0.002 | 9.13 | 5 | additive |
| 16p13.11_dup | ASD | pgs_asd | 0.87 | 0.03 | 6 | interactive |
| 16p13.11_dup | ADHD | pgs_adhd | 0.0031 | 8.77 | 4 | cnv_only |
| 16p13.11_dup | ADHD | pgs_adhd | <2.2E-16 | 945.19 | 4 | pgs_only |
| 16p13.11_dup | ADHD | pgs_adhd | 0.0045 | 8.08 | 5 | additive |
| 16p13.11_dup | ADHD | pgs_adhd | 0.0056 | 7.66 | 6 | interactive |
| 16p13.11_dup | SSD | pgs_scz | 0.03 | 4.4 | 4 | cnv_only |
| 16p13.11_dup | SSD | pgs_scz | <2.2E-16 | 560.36 | 4 | pgs_only |
| 16p13.11_dup | SSD | pgs_scz | 0.04 | 4.17 | 5 | additive |
| 16p13.11_dup | SSD | pgs_scz | 0.88 | 0.02 | 6 | interactive |
| 16p13.11_dup | MDD | pgs_mdd | 0.09 | 2.94 | 4 | cnv_only |
| 16p13.11_dup | MDD | pgs_mdd | <2.2E-16 | 1484.8 | 4 | pgs_only |
| 16p12.1_del | ASD | pgs_asd | 0.68 | 0.17 | 4 | cnv_only |
| 16p12.1_del | ASD | pgs_asd | <2.2E-16 | 199.99 | 4 | pgs_only |
| 16p12.1_del | ASD | pgs_asd | 0.69 | 0.18 | 5 | additive |
| 16p12.1_del | ASD | pgs_asd | 0.60 | 0.28 | 6 | interactive |
| 16p12.1_del | ADHD | pgs_adhd | 0.027 | 4.91 | 4 | cnv_only |
| 16p12.1_del | ADHD | pgs_adhd | <2.2E-16 | 945.19 | 4 | pgs_only |
| 16p12.1_del | ADHD | pgs_adhd | 0.03 | 4.53 | 5 | additive |
| 16p12.1_del | ADHD | pgs_adhd | 0.66 | 0.19 | 6 | interactive |
| 16p12.1_del | SSD | pgs_scz | 0.82 | 0.05 | 4 | cnv_only |
| 16p12.1_del | SSD | pgs_scz | <2.2E-16 | 560.36 | 4 | pgs_only |
| 16p12.1_del | SSD | pgs_scz | 0.70 | 0.15 | 5 | additive |
| 16p12.1_del | SSD | pgs_scz | 0.06 | 3.39 | 6 | interactive |
| 16p12.1_del | MDD | pgs_mdd | 0.34 | 0.92 | 4 | cnv_only |
| 16p12.1_del | MDD | pgs_mdd | <2.2E-16 | 1484.8 | 4 | pgs_only |
| 16p11.2_dup | ASD | pgs_asd | 9.96E-06 | 19.52 | 4 | cnv_only |
| 16p11.2_dup | ASD | pgs_asd | <2.2E-16 | 199.99 | 4 | pgs_only |
| 16p11.2_dup | ASD | pgs_asd | 9.66E-06 | 19.58 | 5 | additive |
| 16p11.2_dup | ASD | pgs_asd | 0.96 | 0 | 6 | interactive |
| 16p11.2_dup | ADHD | pgs_adhd | 7.58E-09 | 33.38 | 4 | cnv_only |
| 16p11.2_dup | ADHD | pgs_adhd | <2.2E-16 | 945.19 | 4 | pgs_only |
| 16p11.2_dup | ADHD | pgs_adhd | 2.95E-09 | 35.22 | 5 | additive |
| 16p11.2_dup | ADHD | pgs_adhd | 0.76 | 0.09 | 6 | interactive |
| 16p11.2_dup | SSD | pgs_scz | 0.02 | 5.38 | 4 | cnv_only |
| 16p11.2_dup | SSD | pgs_scz | <2.2E-16 | 560.36 | 4 | pgs_only |
| 16p11.2_dup | SSD | pgs_scz | 0.02 | 5.45 | 5 | additive |
| 16p11.2_dup | SSD | pgs_scz | 0.78 | 0.08 | 6 | interactive |
| 16p11.2_dup | MDD | pgs_mdd | 0.53 | 0.4 | 4 | cnv_only |
| 16p11.2_dup | MDD | pgs_mdd | <2.2E-16 | 1484.8 | 4 | pgs_only |
| 22q11.2_dup | ASD | pgs_asd | 0.0019 | 9.65 | 4 | cnv_only |
| 22q11.2_dup | ASD | pgs_asd | <2.2E-16 | 199.99 | 4 | pgs_only |
| 22q11.2_dup | ASD | pgs_asd | 0.0016 | 9.99 | 5 | additive |
| 22q11.2_dup | ASD | pgs_asd | 0.29 | 1.14 | 6 | interactive |
| 22q11.2_dup | ADHD | pgs_adhd | 0.0008 | 11.23 | 4 | cnv_only |
| 22q11.2_dup | ADHD | pgs_adhd | <2.2E-16 | 945.19 | 4 | pgs_only |
| 22q11.2_dup | ADHD | pgs_adhd | 0.00046 | 12.27 | 5 | additive |
| 22q11.2_dup | ADHD | pgs_adhd | 0.91 | 0.01 | 6 | interactive |
| 22q11.2_dup | SSD | pgs_scz | 0.26 | 1.26 | 4 | cnv_only |
| 22q11.2_dup | SSD | pgs_scz | <2.2E-16 | 560.36 | 4 | pgs_only |
| 22q11.2_dup | SSD | pgs_scz | 0.16 | 2 | 5 | additive |
| 22q11.2_dup | SSD | pgs_scz | 0.445 | 0.57 | 6 | interactive |
| 22q11.2_dup | MDD | pgs_mdd | 0.81 | 0.06 | 4 | cnv_only |
| 22q11.2_dup | MDD | pgs_mdd | <2.2E-16 | 1484.8 | 4 | pgs_only |

### Supplementary Table 9

*Model fitting results for Schizophrenia as a function of PGS and rCNV status (using rCNVs as LOEUF groups and aggregated).*

| **Model** | **Genetic exposure ^a^** | **df** | **Chisq ^b^** | **P ^c^** | **Delta R^2 d^** |
| --- | --- | --- | --- | --- | --- |
| 0 | None | 3 |  |  |  |
| 1 | CNV_groups | 4 | 27.42 | 4.81 x 10^-6^ | 0.0013 |
| 2 | anyCNV | 4 | 21.21 | 4.12 x 10^-6^ | 0.0001 |
| 3 | PGS | 4 | 386.13 | <2.2 x 10^-16^ | 0.0179 |
| 4 | PGS + CNV_groups | 5 | 25.41 | 1.27 e-5 | 0.0191 |
| 5 | PGS + anyCNV | 5 | 384.41 | <2.2 x 10^-16^ | 0.0188 |
| 6 | PGS×CNV_groups | 8 | 3.80 | 0.28 | 0.0192 |
| 7 | PGS×anyCNV | 6 | 2.12 | 0.14 | 0.0189 |

*In addition to the main analysis of assessing the joint effect of rCNV and PGS on schizophrenia spectrum disorder (SSD; ICD10:F20-29) by GLMs, we performed similar analyses using the narrow definition for schizophrenia (SCZ; ICD10: F20) instead (see* ***Methods*** *and* ***sTable 5a*** *for more details). Likelihood ratio tests were used to compare each model with its nested model. The null model included sex, age at the end of follow-up, and the genotyping array as independent variables. rCNV and PGS were added in a stepwise manner to the null model. ^a^ “anyCNV” and “CNV_groups” represent rCNVs in aggregated form and rCNV LOEUF-groups, respectively .^b^ Chisq and ^c^ P are derived from the likelihood ratio tests. ^d^ Nagelkerke’s R2 was calculated for each model and ΔR^2^ was derived by subtracting the R2 of each full model from R2 of the subsequent nested model.*

### Supplementary Table 10

*CNV×PRS Interaction on ASD, ADHD, and SSD for rCNV-LOEUF groups, aggregated rCNVs, and common individual rCNVs.*

|  | ***ASD*** | | | | ***ADHD*** | | | ***SSD*** | | |
| --- | --- | --- | --- | --- | --- | --- | --- | --- | --- | --- |
|  | ***CNV× ASD-PGS*** | | | | ***CNV× ADHD-PGS*** | | | ***CNV× SCZ-PGS*** | | |
|  | | ***β*** | ***SE*** | ***p*** | ***β*** | ***SE*** | ***P*** | ***β*** | ***SE*** | ***P*** |
| ***Low LOEUF*** | | *0.01* | *0.08* | *0.93* | *-0.06* | *0.07* | *0.40* | *-0.05* | *0.09* | *0.60* |
| ***Medium LOEUF*** | | *-0.16* | *0.11* | *0.14* | *-0.15* | *0.11* | *0.18* | *-0.2* | *0.12* | *0.11* |
| ***High LOEUF*** | | *-0.01* | *0.13* | *0.95* | *0.12* | *0.13* | *0.38* | *0.14* | *0.17* | *0.41* |
| ***Any CNV*** | | *-0.04* | *0.06* | *0.48* | *-0.06* | *0.05* | *0.29* | *-0.06* | *0.07* | *0.34* |
| ***TAR _dup*** | | *-0.31* | *0.29* | *0.29* | *-0.03* | *0.27* | *0.91* | *0.23* | *0.30* | *0.46* |
| ***1q21.1_dup*** | | *-0.16* | *0.30* | *0.60* | *-0.11* | *0.25* | *0.66* | *-0.53* | *0.28* | *0.06* |
| ***15q11.2_del*** | | *-0.15* | *0.28* | *0.60* | *0.03* | *0.27* | *0.91* | *-0.44* | *0.37* | *0.24* |
| ***15q11.2_dup*** | | *0.05* | *0.30* | *0.86* | *0.33* | *0.35* | *0.34* | *-0.02* | *0.35* | *0.96* |
| ***16p13.11_del*** | | *-0.10* | *0.28* | *0.73* | *0.28* | *0.34* | *0.42* | *-0.47* | *0.34* | *0.17* |
| ***16p13.11_dup*** | | *-0.03* | *0.18* | *0.87* | *-0.51* | *0.18* | ***0.0057*** | *0.04* | *0.25* | *0.88* |
| ***16p12.1_del*** | | *0.18* | *0.16* | *0.28* | *-0.07* | *0.12* | *0.58* | *0.03* | *0.17* | *0.84* |
| ***16p11.2_dup*** | | *0.01* | *0.14* | *0.95* | *-0.09* | *0.13* | *0.46* | *-0.07* | *0.18* | *0.71* |
| ***22q11.2_dup*** | | *-0.01* | *0.27* | *0.96* | *-0.07* | *0.23* | *0.76* | *-0.09* | *0.32* | *0.78* |

*Interaction terms are attributed to the fitted generalized linear models (GLMs) for assessing the interactive effects of rCNV and PGS on ASD, ADHD, and SSD (See method, sTable 5a and sTable 5b). We tested rCNV×PGS* *using rCNVs groups (divided based on their LOEUF scores), common individual rCNVs, and aggregated rCNVs (any rCNV vs. no rCNV). Interaction effects on MDD are not reported since we found no indication of significant effect of rCNVs on MDD.*

### Supplementary Table 11

*Number of carriers of the common rCNVs in iPSYCH2015 across cohort and case samples (Unrelated European sample)*

| **rCNV** | **iPSYCH^a^** | **Cohort^b^** | **ASD** | **ADHD** | **SSD** | **MDD** |
| --- | --- | --- | --- | --- | --- | --- |
| **TAR_dup** | 131 | 42 | 26 | 40 | 14 | 37 |
| **1q21.1_dup** | 103 | 26 | 41 | 32 | 27 | 17 |
| **15q11.2_del** | 484 | 155 | 114 | 123 | 40 | 123 |
| **15q11_dup** | 448 | 157 | 72 | 110 | 34 | 116 |
| **16p13.11_del** | 105 | 27 | 22 | 32 | 13 | 20 |
| **16p1311_dup** | 226 | 66 | 55 | 59 | 19 | 58 |
| **16p12.1_del** | 128 | 45 | 21 | 36 | 19 | 30 |
| **16p11.2_dup** | 146 | 34 | 39 | 61 | 48 | 33 |
| **22q11_dup** | 127 | 37 | 35 | 42 | 13 | 24 |

*In addition to the main analyses of assessing the joint effects of PGS and rCNVs (i.e., in the form of LOEUF groups and aggregated) on ASD, ADHD, SSD, and MDD using GLM models, we conducted similar GLM analyses using common individual rCNVs that had over* $1/1000$ *carriers in the entire sample. ^a^ Some individuals are diagnosed with more than one condition, therefore the number of rCNV carriers in the entire iPSYCH does not correspond to the sum of the carrier counts across columns of cases and cohort. ^b^ Due to the overlap between cases and the random cohort, some carriers within the random cohort may be part of case samples too.*

### Supplementary Table 12

*Average PGS for carriers and non-carriers across ASD, ADHD, SSD, and MDD by affection status*

|  | ***ASD*** | | | | ***ADHD*** | | | |
| --- | --- | --- | --- | --- | --- | --- | --- | --- |
|  | ***Cases*** | | ***Controls*** | | ***Cases*** | | ***Controls*** | |
| ***CNV group*** | ***Mean*** | ***SE*** | ***Mean*** | ***SE*** | ***Mean*** | ***SE*** | ***Mean*** | ***SE*** |
| *No CNV* | *0.11* | *0.008* | *-0.04* | *0.04* | *0.18* | *0.08* | *-0.10* | *0.005* |
| *Low LOEUF* | *0.09* | *0.05* | *-0.05* | *0.07* | *0.14* | *0.05* | *-0.11* | *0.04* |
| *Medium LOEUF* | *-0.04* | *0.09* | *-0.06* | *0.09* | *0.09* | *0.07* | *-0.02* | *0.07* |
| *High LOEUF* | *0.08* | *0.08* | *-0.01* | *0.03* | *0.01* | *0.07* | *-0.25* | *0.08* |
| *Any CNV* | *0.06* | *0.040* | *-0.04* | *0.005* | *0.12* | *0.04* | *-0.11* | *0.03* |
|  | ***SSD*** | | | | ***MDD*** | | | |
| *No CNV* | *0.21* | *0.01* | *-0.06* | *0.005* | *0.16* | *0.006* | *-0.17* | *0.005* |
| *Low LOEUF* | *0.25* | *0.08* | *-0.08* | *0.05* | *0.20* | *0.05* | *-0.15* | *0.04* |
| *Medium LOEUF* | *0.15* | *0.11* | *0.05* | *0.08* | *0.15* | *0.08* | *-0.20* | *0.08* |
| *High LOEUF* | *0.24* | *0.14* | *-0.15* | *0.09* | *0.29* | *0.11* | *-0.2* | *0.09* |
| *Any CNV* | *0.22* | *0.06* | *-0.06* | *0.04* | *0.20* | *0.04* | *-0.17* | *0.04* |

*The Mean of PGS corresponding to each disorder was computed across different groups of individuals (i.e., no rCNV, any rCNV, and rCNV LOEUF-groups) among cases and controls for each disorder separately. The differences in average PGS between the groups across the four diagnoses were assessed using Welsch’s t-test (see* ***sTable 9****).*

### Supplementary Table 13

*Welsch’s T-test results for comparison of average PGS among rCNV carriers of LOEUF groups and non-carriers across ASD, ADHD, SSD, and MDD. p_cases and p_controls attribute to the p values derive from comparisons among cases and controls,*

*separately. ADHD; attention-deficit hyperactivity disorder, ASD; autism spectrum disorder, MDD; major depressive disorder, SSD; schizophrenia spectrum disorder. All the P-values were adjusted by applying FDR correction with p_adjust() function in R.*

| **Diagnosis** | **PGS** | **Comparison test** | **p_cases** | **p_controls** |
| --- | --- | --- | --- | --- |
| ADHD | pgs_adhd | Medium LOEUF vs Low LOEUF | 0.78 | 0.51 |
| ADHD | pgs_adhd | Medium LOEUF vs High LOEUF | 0.97 | 0.36 |
| ADHD | pgs_adhd | Medium LOEUF vs noCNV | 0.51 | 0.51 |
| ADHD | pgs_adhd | Low LOEUF vs noCNV | 0.68 | 0.92 |
| ADHD | pgs_adhd | High LOEUF vs Low LOEUF | 0.78 | 0.48 |
| ADHD | pgs_adhd | High LOEUF vs noCNV | 0.51 | 0.36 |
| ASD | pgs_asd | Medium LOEUF vs Low LOEUF | 0.93 | 0.93 |
| ASD | pgs_asd | Medium LOEUF vs High LOEUF | 0.93 | 0.93 |
| ASD | pgs_asd | Medium LOEUF vs noCNV | 0.93 | 0.93 |
| ASD | pgs_asd | Low LOEUF vs noCNV | 0.93 | 0.93 |
| ASD | pgs_asd | High LOEUF vs Low LOEUF | 0.93 | 0.93 |
| ASD | pgs_asd | High LOEUF vs noCNV | 0.93 | 0.93 |
| SSD | pgs_scz | Medium LOEUF vs Low LOEUF | 0.86 | 0.38 |
| SSD | pgs_scz | Medium LOEUF vs High LOEUF | 0.86 | 0.38 |
| SSD | pgs_scz | Medium LOEUF vs noCNV | 0.86 | 0.38 |
| SSD | pgs_scz | Low LOEUF vs noCNV | 0.86 | 0.99 |
| SSD | pgs_scz | High LOEUF vs Low LOEUF | 0.99 | 0.86 |
| SSD | pgs_scz | High LOEUF vs noCNV | 0.98 | 0.86 |
| MDD | pgs_mdd | Medium LOEUF vs Low LOEUF | 0.93 | 0.93 |
| MDD | pgs_mdd | Medium LOEUF vs High LOEUF | 0.93 | 0.93 |
| MDD | pgs_mdd | Medium LOEUF vs noCNV | 0.93 | 0.93 |
| MDD | pgs_mdd | Low LOEUF vs noCNV | 0.93 | 0.93 |
| MDD | pgs_mdd | High LOEUF vs Low LOEUF | 0.93 | 0.99 |
| MDD | pgs_mdd | High LOEUF vs noCNV | 0.93 | 0.93 |
